# ^18^F-NaF Uptake on Vascular PET Imaging in Symptomatic versus Asymptomatic Atherosclerotic Disease: a Meta-Analysis

**DOI:** 10.1101/2024.02.22.24303229

**Authors:** S Bhakta, MM Chowdhury, JM Tarkin, JHF Rudd, EA Warburton, NR Evans

**Affiliations:** Department of Clinical Neurosciences, University of Cambridge, Cambridge. CB2 0QQ; Department of Vascular Surgery, University of Cambridge, Cambridge. CB2 0QQ; Division of Cardiovascular Medicine, University of Cambridge, Cambridge. CB2 0QQ

## Abstract

**Introduction:** ^18^F-sodium fluoride (NaF) positron-emission tomography (PET) is increasingly being used to measure microcalcification in atherosclerotic disease *in vivo*. Correlations have been drawn between sodium fluoride uptake and the presence of high-risk plaque features, as well as its association with clinical atherosclerotic sequelae. The aim of this review was to perform a meta-analysis of NaF uptake on PET imaging and its relation to symptomatic and asymptomatic disease.

**Methods:** A systematic review was performed according to PRISMA guidelines, via searching the MEDLINE database up to August 2023. The search strategy included the terms “NaF”, “PET” and “plaque”, and all studies were included where there was data listed regarding the degree of microcalcification, as measured by ^18^F-NaF uptake in symptomatic and asymptomatic atherosclerotic plaques. Analysis involved calculating standardized mean differences between uptake values and comparison using a random-effects model.

**Results:** A total of 15 articles, involving 423 participants, were included in the meta-analysis. Comparing ^18^F-NaF uptake in symptomatic vs asymptomatic atherosclerotic plaques, a standardized mean difference of 0.42 (95% CI 0.29-0.56; p<0.001, I^2^ = 54.1%) was noted for those studies comparing symptomatic and asymptomatic plaques in the same participant, with no significant change in effect based on arterial territory studied (Q_M_ = 5.02, p = 0.08). In those studies where data was included from participants with and without symptomatic disease, the standardized mean difference between symptomatic and asymptomatic plaques was 0.44 (95% CI 0.03-0.85, p=0.037, I^2^ = 40.4%). All studies including asymptomatic participants were investigating carotid disease.

**Conclusions:** PET imaging using ^18^F-NaF can detect differences in microcalcification between symptomatic and asymptomatic atherosclerotic plaques within and between individuals, and is a marker of symptomatic disease. The standardization of ^18^F-NaF PET imaging protocols, and its future use as a risk stratification tool or outcome measure, requires further study.

## Introduction

Atherosclerosis is a systemic chronic arterial disease^1^, involving the accumulation of lipids and inflammatory cells^2^ to form foci of disease, termed ‘plaques’, at the vessel wall. It is the cause of over one third of all deaths^3^, through resulting diseases such as myocardial infarction, ischemic stroke, and critical limb threatening ischemia. In some patients, minor or less severe clinical symptoms may be a marker of higher risk for progressing to more severe clinical disease, such as stable angina preceding myocardial infarction^4,5^, or transient ischemic attack, conferring a higher risk of ischemic stroke in the short term^6,7^.

Plaques may have heterogenous appearances^8^, with certain plaque features indicating a higher risk of rupture and subsequent clinical sequelae^8,9^. These high-risk features include the presence of a lipid-rich necrotic core^10^, intraplaque hemorrhage^11^, a thin or ruptured fibrous cap^12^, and the presence of microcalcification^13^. The mechanisms determining the transition from a low-risk (“stable”) to a high-risk (“unstable”) plaque and vice versa are incompletely understood^14,15^, but microcalcification has been recognized as a potential cause for acute plaque rupture, through mechanical destabilization of the fibrous cap of the plaque^16^, as well as causing an increased inflammatory response within the plaque^17^, leading to enzymatic destabilization of the plaque^18^. Microcalcifications are calcium deposits of <50μm in diameter, which is below the spatial resolution of commonly-used clinical vascular imaging techniques^19^, such as computed tomography (CT) or magnetic resonance imaging (MRI).

Positron emission tomography (PET) is a nuclear imaging technique used in vascular imaging due to its high sensitivity to detect low concentrations of radiolabeled ligands (termed “tracers”), which can be directed to detect the presence of a specific target or process^20^. Sodium fluoride (NaF), labelled with fluorine-18, has been validated as a tracer for the identification of microcalcification in vascular imaging^21^.

NaF adsorbs to the surface of hydroxyapatite within the body^22^, with hydroxyapatite being the most common calcium-containing crystal structure in atherosclerotic plaques *in vivo*^23,24^. Fluorine exchanges with hydroxyl groups on the surface of these crystals, while there is substantially less tracer uptake deeper within the crystal structure of the molecule^22^. Calcium deposits with an increased surface area therefore have increased fluoride ion uptake over the uptake durations used in clinical PET scanning^22,25^. Using NaF, this process of hydroxyl-ion substitution can be used to detect microcalcifications via PET imaging. Due to their high surface area to volume ratio, microcalcifications will demonstrate higher uptake on PET compared to areas with no microcalcification, or those with larger deposits of calcium (“macrocalcification”)^26–28^. NaF-PET can be preferred for vascular imaging compared to other PET imaging techniques for visualizing high-risk plaques due to issues, such as ‘spill-over artefact’ when using fluorodeoxyglucose (FDG)-PET in cardiac imaging.

There is an increasing body of literature demonstrating the use of NaF-PET for vascular imaging in atherosclerosis^29^, and specifically, in symptomatic disease. This meta-analysis focuses on the role of NaF-PET imaging to differentiate between symptomatic versus asymptomatic atherosclerotic disease.

## Methods

### Protocol, Search Strategy and Selection Criteria

Details for the protocol were registered on PROSPERO^30^. The selection process and reporting items were based on the preferred reporting items for systematic reviews and meta-analysis (PRISMA) flow diagram and checklist^31^. The primary outcome was to determine differences in NaF tracer uptake between symptomatic and asymptomatic atherosclerotic plaques. A search strategy was formulated using Embase and Medline All via Ovid (**Figure 1**). Additionally, a manual search was performed to identify relevant records through reference searches. Duplicate records were removed, and the retrieved records were checked for inclusion and exclusion criteria.

**Figure 1.**
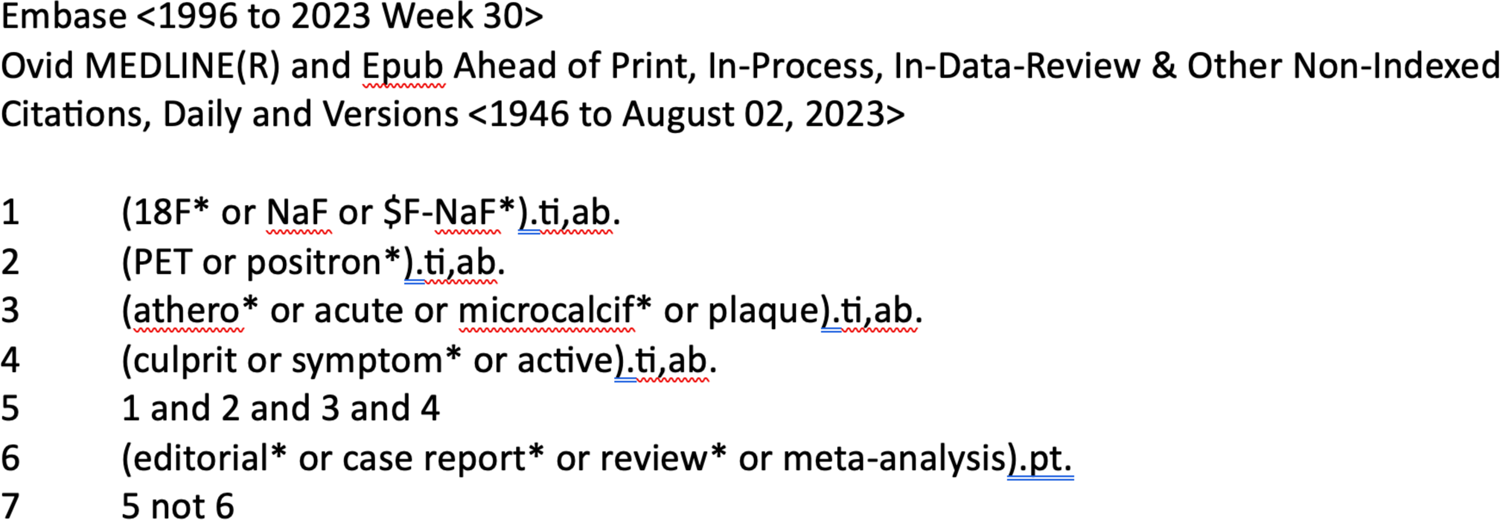
Search strategy performed for systematic review and meta-analysis

Studies were eligible for inclusion if they investigated the association between NaF uptake and symptomatic and asymptomatic atherosclerotic plaques, where symptomatic plaques were those that were associated with a recent clinical vascular event, including, but not limited to, stroke or myocardial infarction, while asymptomatic plaques were those not associated with a recent clinical vascular event. Studies were excluded if animal data was used, non-atherosclerotic disease was investigated, or if the study did not provide details regarding the type or location of symptomatic disease.

### Data Selection, Extraction and Quality Assessment

Data were extracted by one study investigator (S. B.) and checked by another researcher (N. R. E.), from the included records, using a standardized electronic data collection form. Discrepancies were resolved by re-extraction or by third-party adjudication (E. A. W.) as required. Retrieved characteristics from the studies included, but was not limited to, the number of subjects, patient population, targeted vascular territory, dose of NaF injected, uptake time, imaging protocols, primary endpoint measures for PET/CT and main findings, which were tabulated as per published guidance^32^. The ROBINS-E tool^33^ was used to assess the quality of the studies included in this review (**Supplementary** Figure 1).

### Statistical Analysis

The mean and associated standard deviation (SD), or the median and the associated interquartile range (IQR) of the measurement of NaF signal were extracted from the included studies. Extracted median and IQR data was converted into mean and SD data in order to calculate a unified outcome^34^. In studies where multiple measures of NaF signal were listed, the value corresponding to the TBR related to the maximum SUV value (TBR_max_) was taken. The absolute difference between the populations when NaF signal is measured using SUV versus TBR is minor, given the low blood pool activity of NaF following an appropriate uptake time^19^.

Studies were classified by their included participants – those with only symptomatic participants, or those also including an asymptomatic control population. The effect estimates from each study were pooled using the inverse-variance weighted method, and random effects models were used for meta-analysis. *I*^2^ statistics were calculated to determine the variability in effect estimate due to between-study heterogeneity. Meta-analyses were performed where there were two or more studies using the same type of included population. Standardized mean differences were calculated due to the variability in reporting outcomes used in the included studies.

Funnel plots (**Figures 2 and 3**) were used to visually assess the symmetry of the studies about the mean effect size, to identify any publication bias. Two-tailed tests were used, and a *p*-value of 0.05 was taken as the limit of statistical significance. Statistical analyses were performed using the *metafor*^35^ and *metamedian*^36^ packages, using R Statistical Software (v4.3.1, R Core Team 2023).

**Figure 2.**
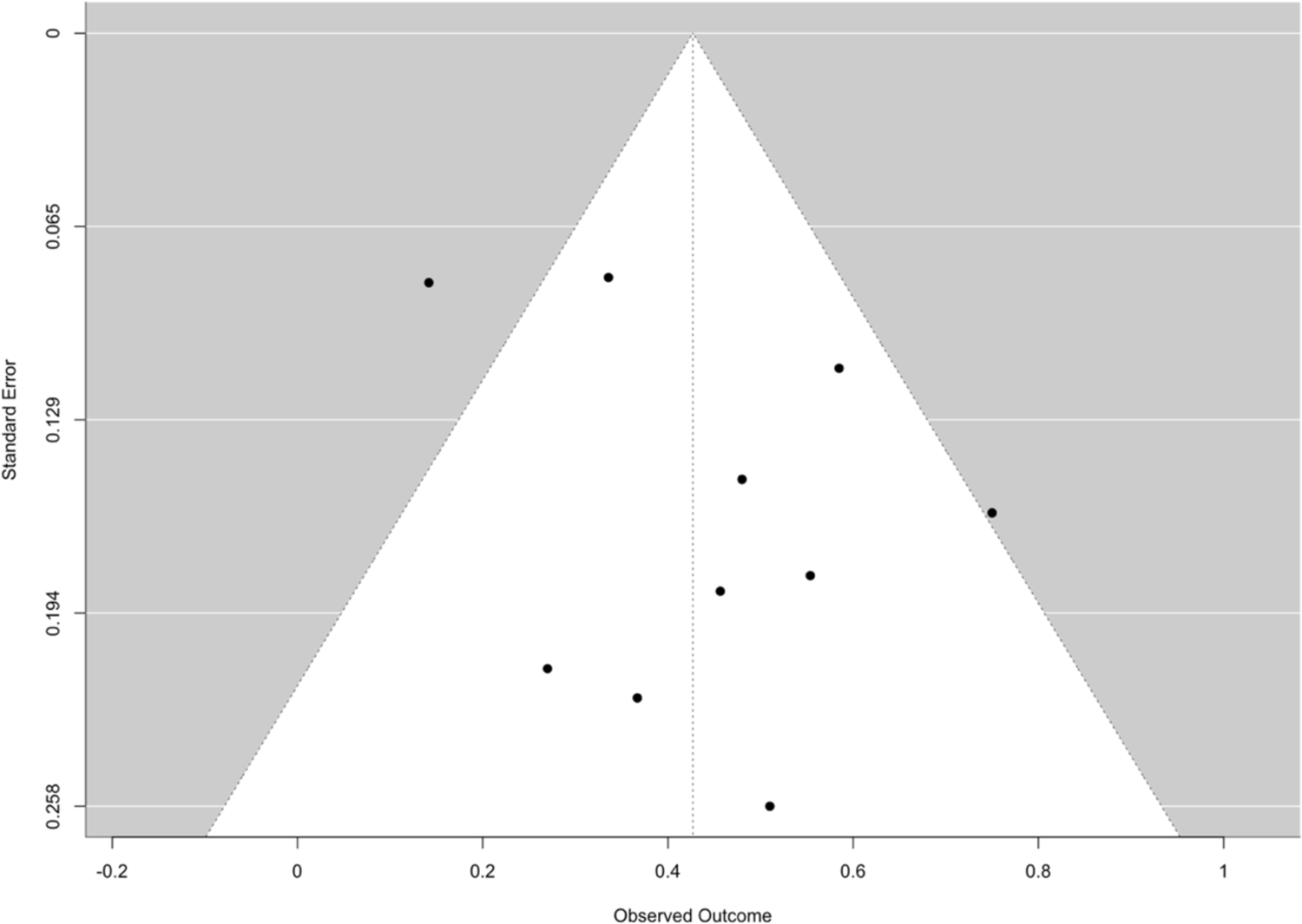
Funnel plot of studies included in Table 1

**Figure 3.**
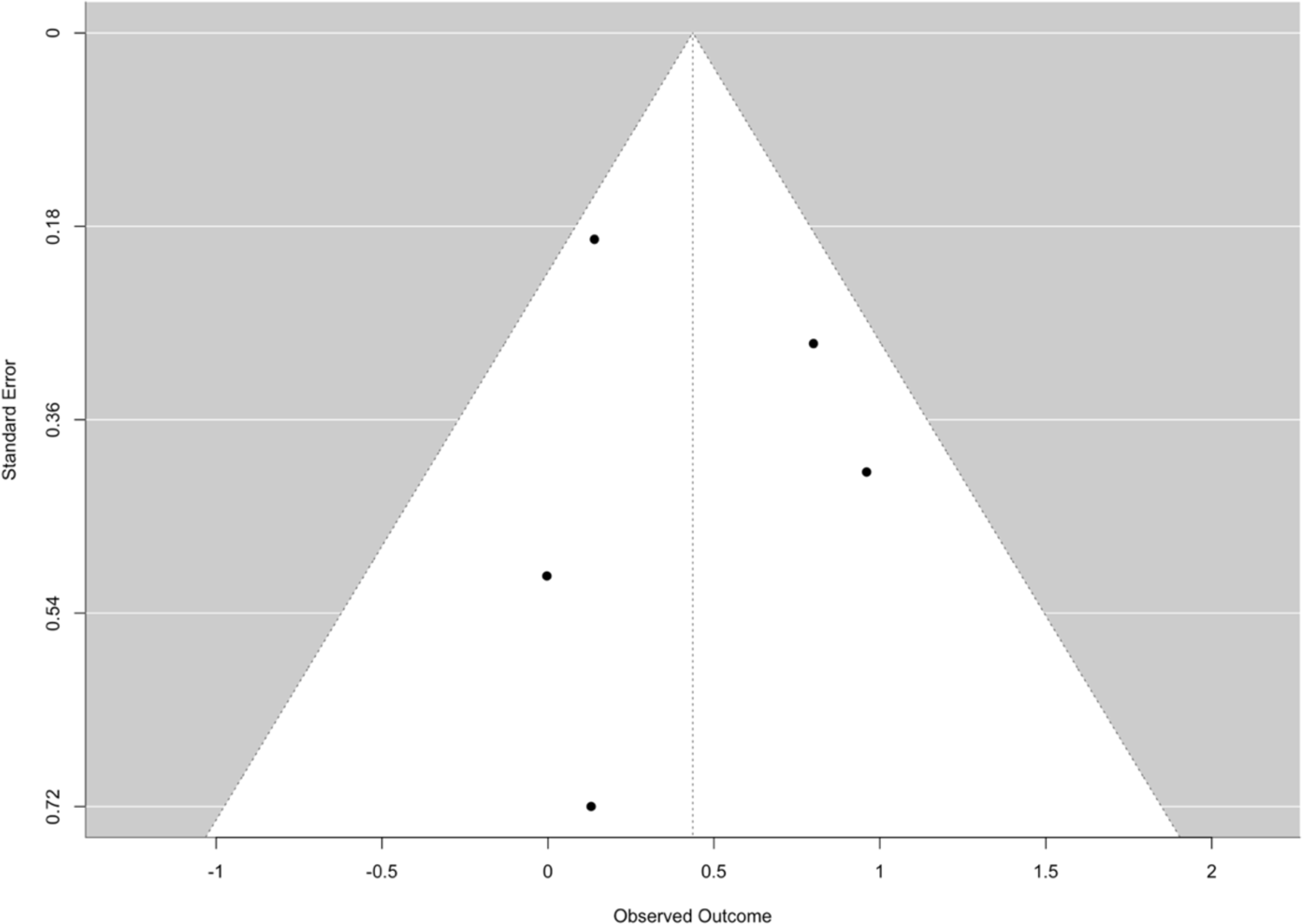
Funnel plot of studies included in Table 2

**Table 1.**
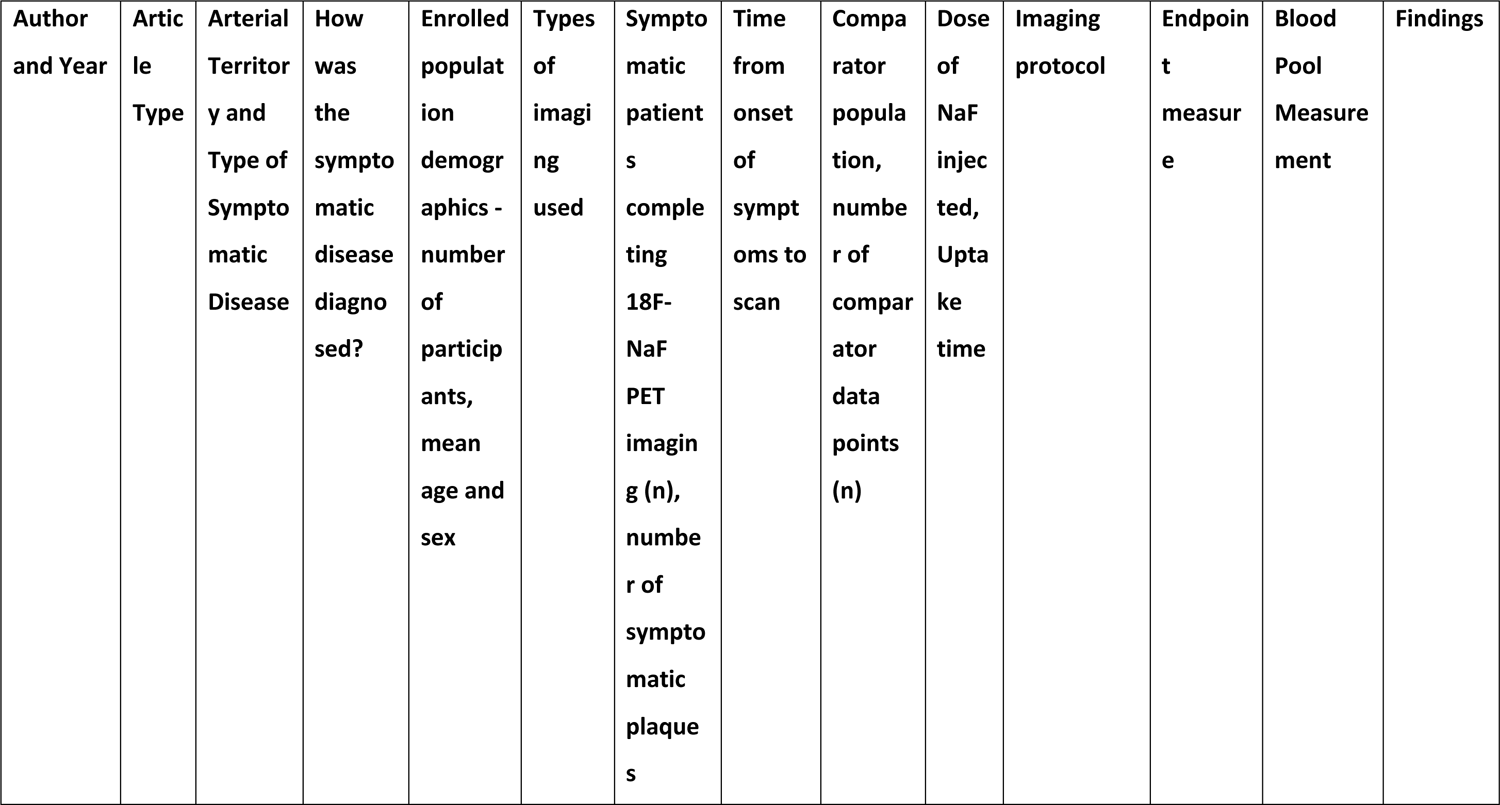
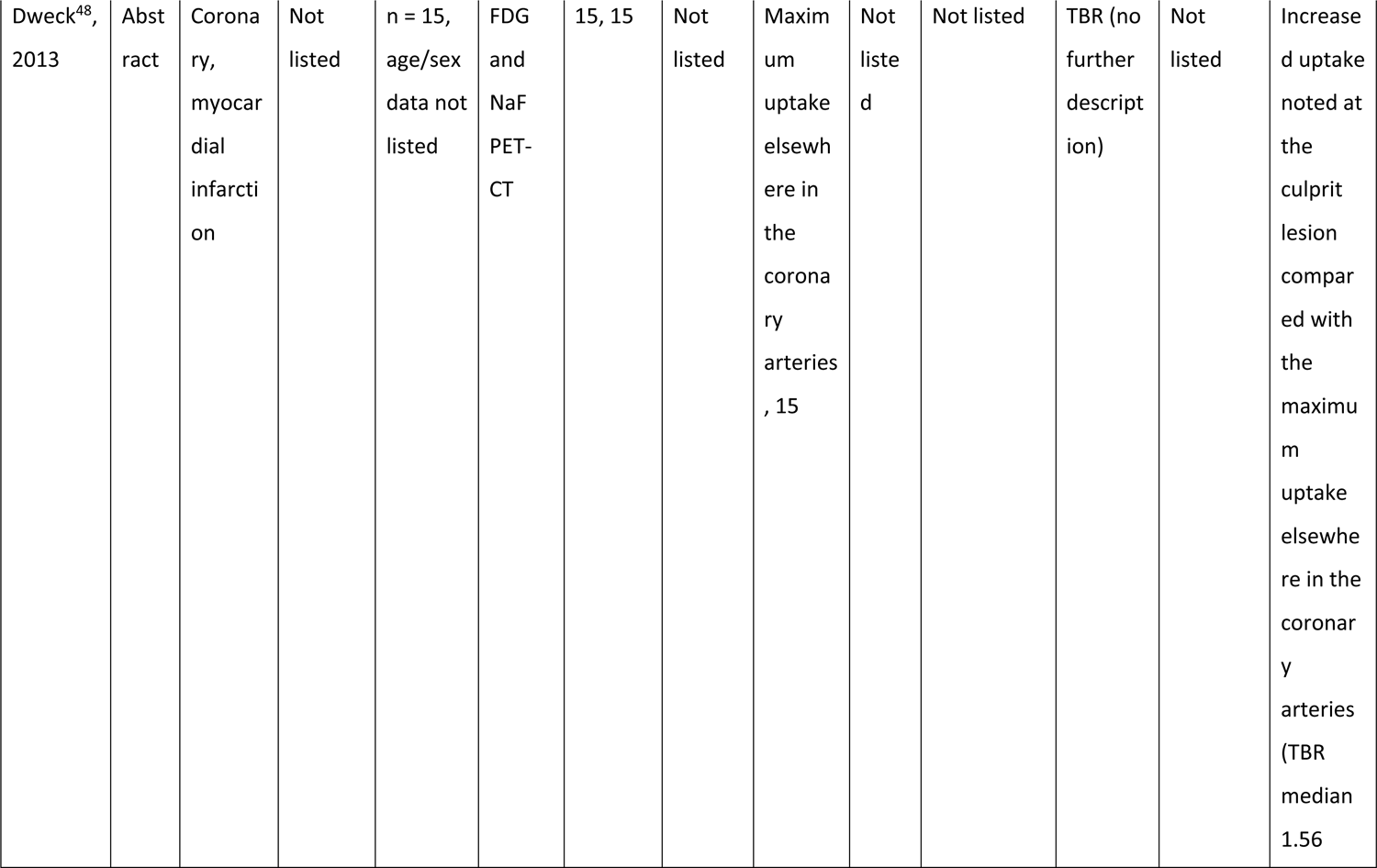
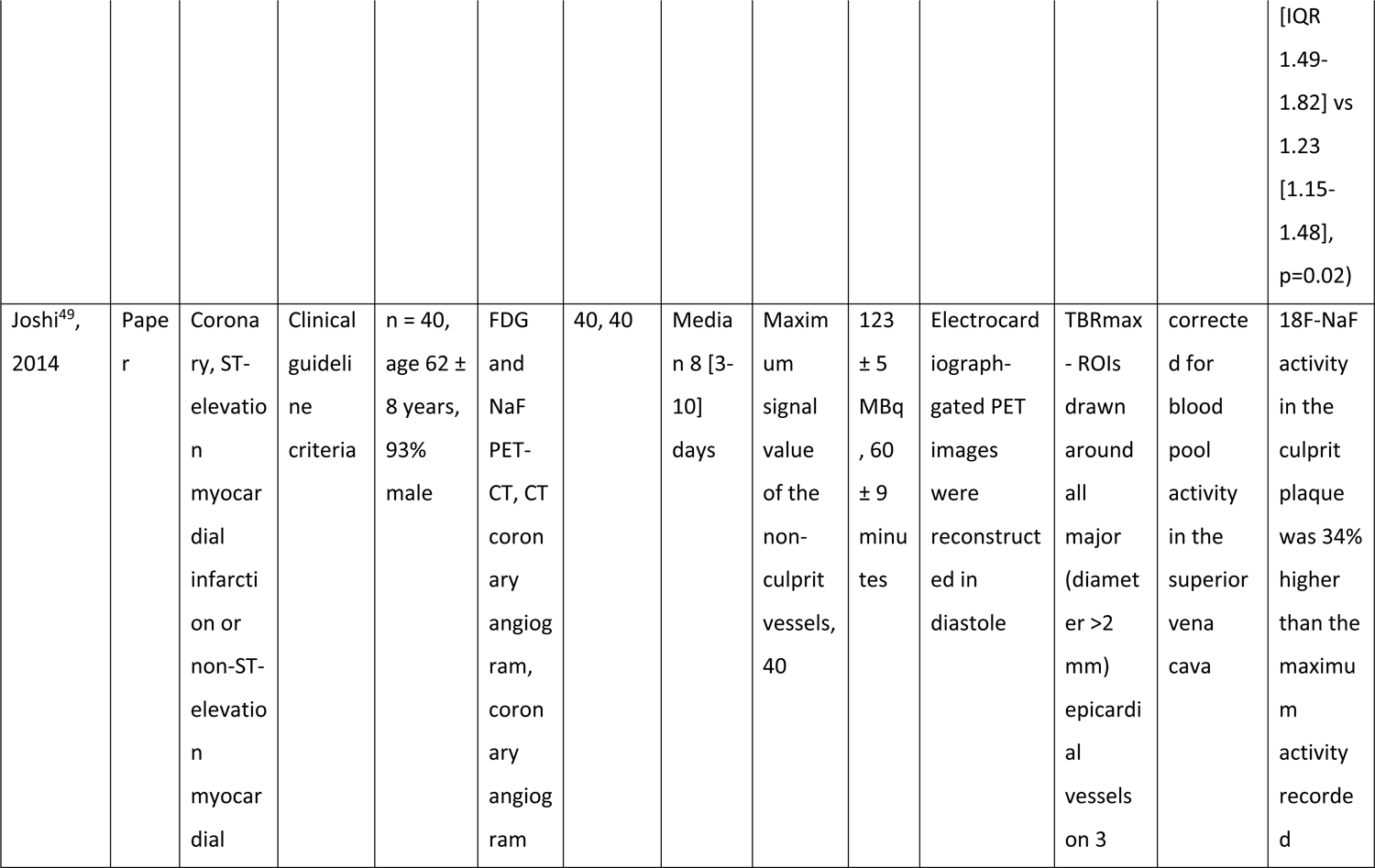
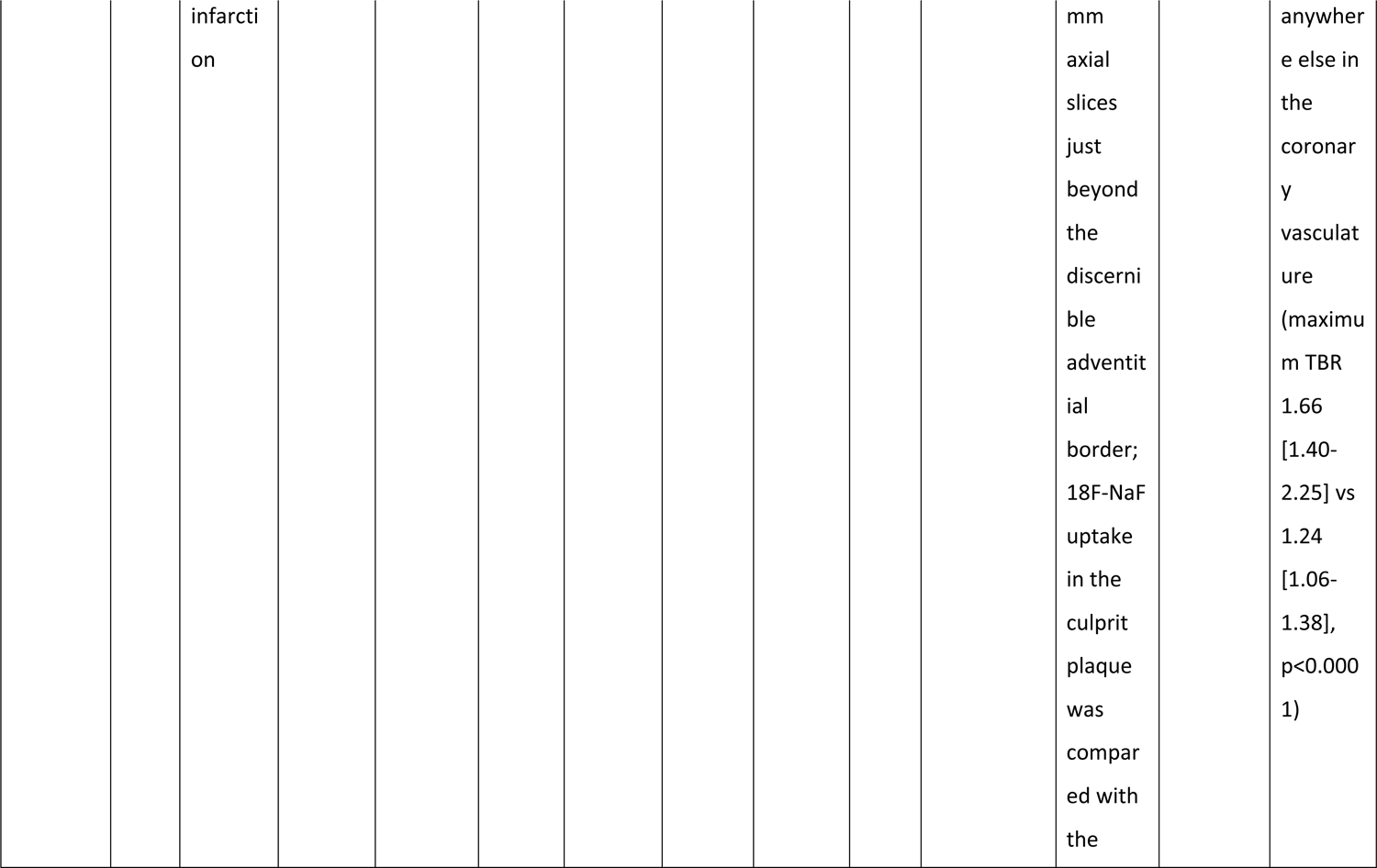
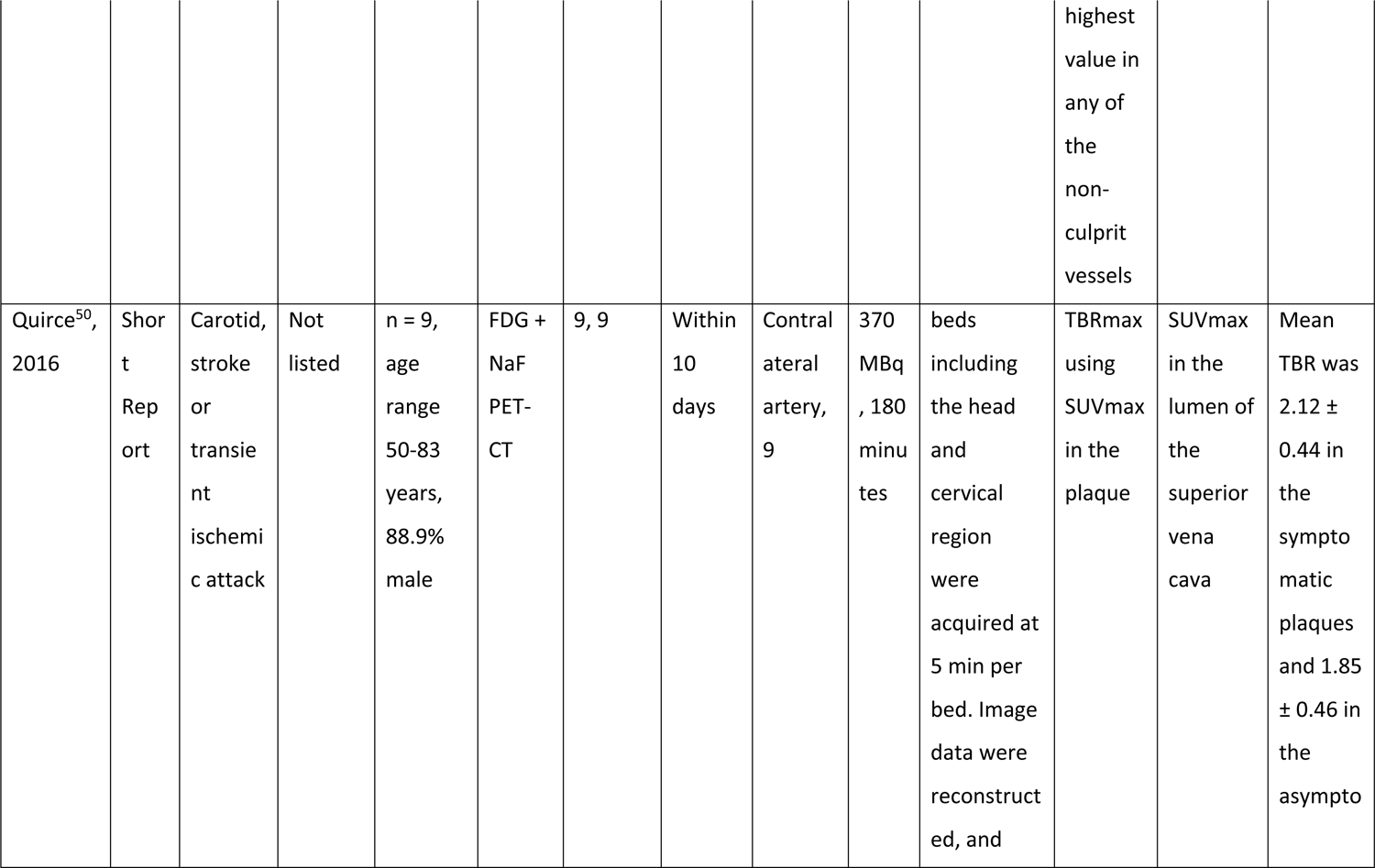
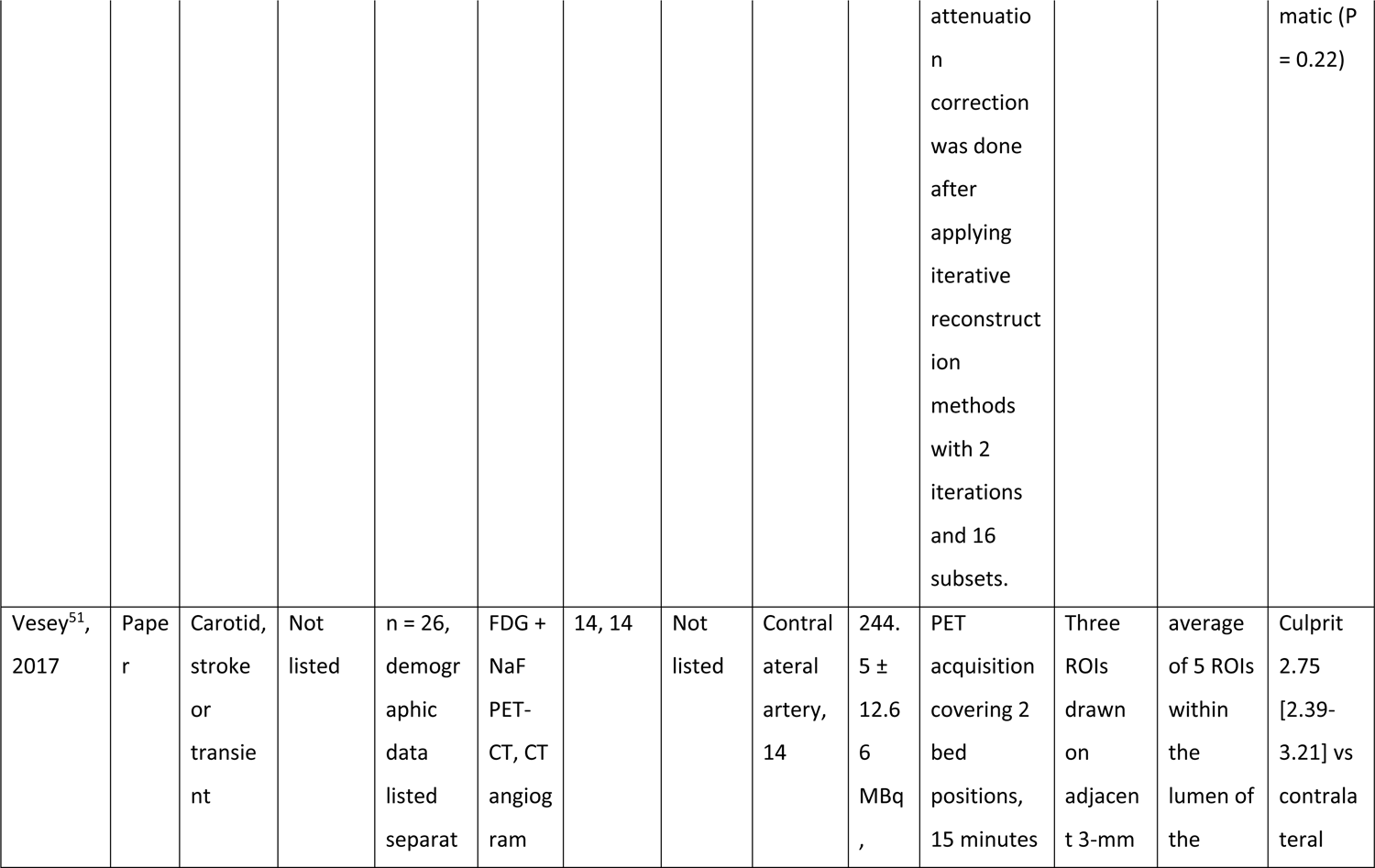
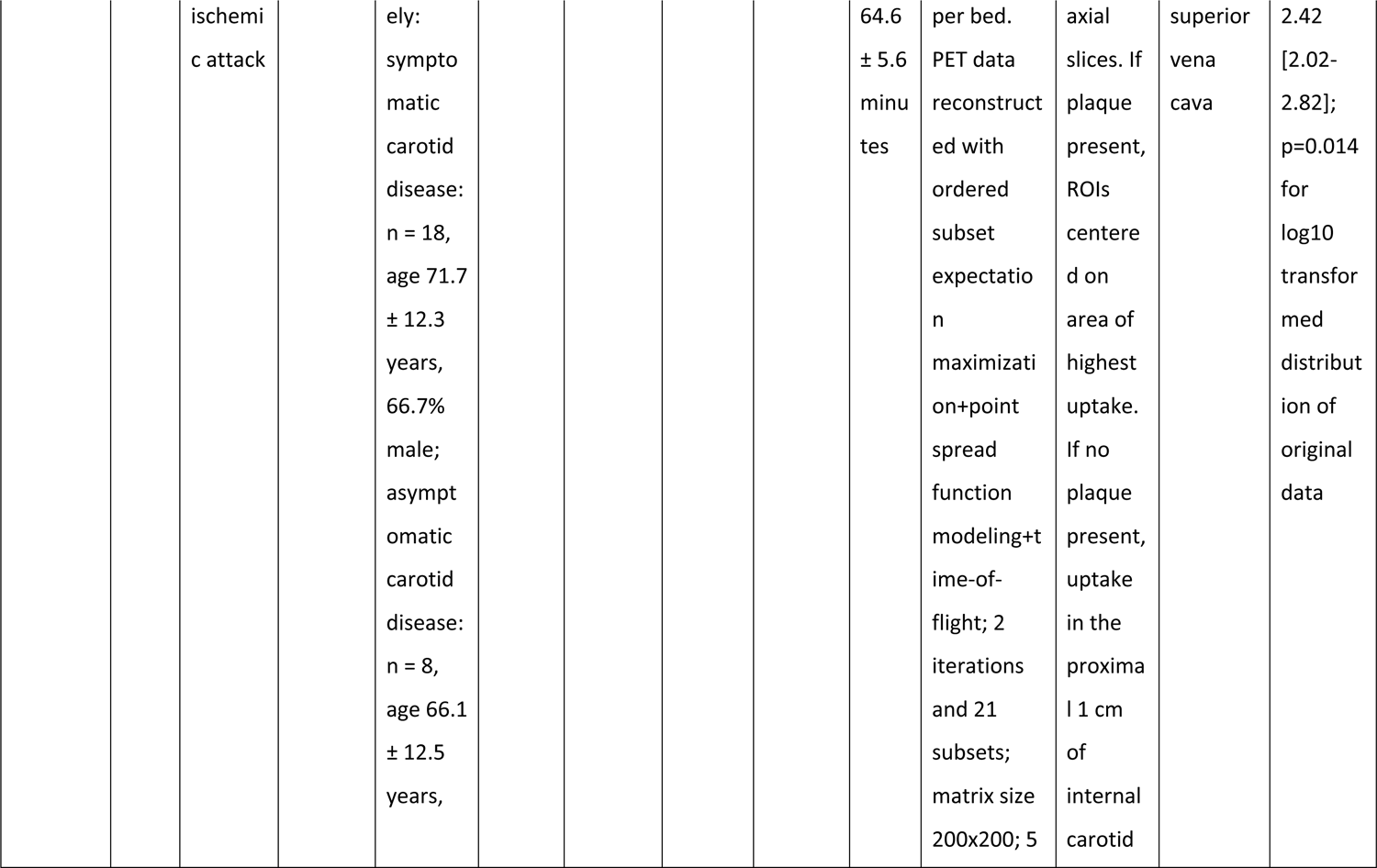
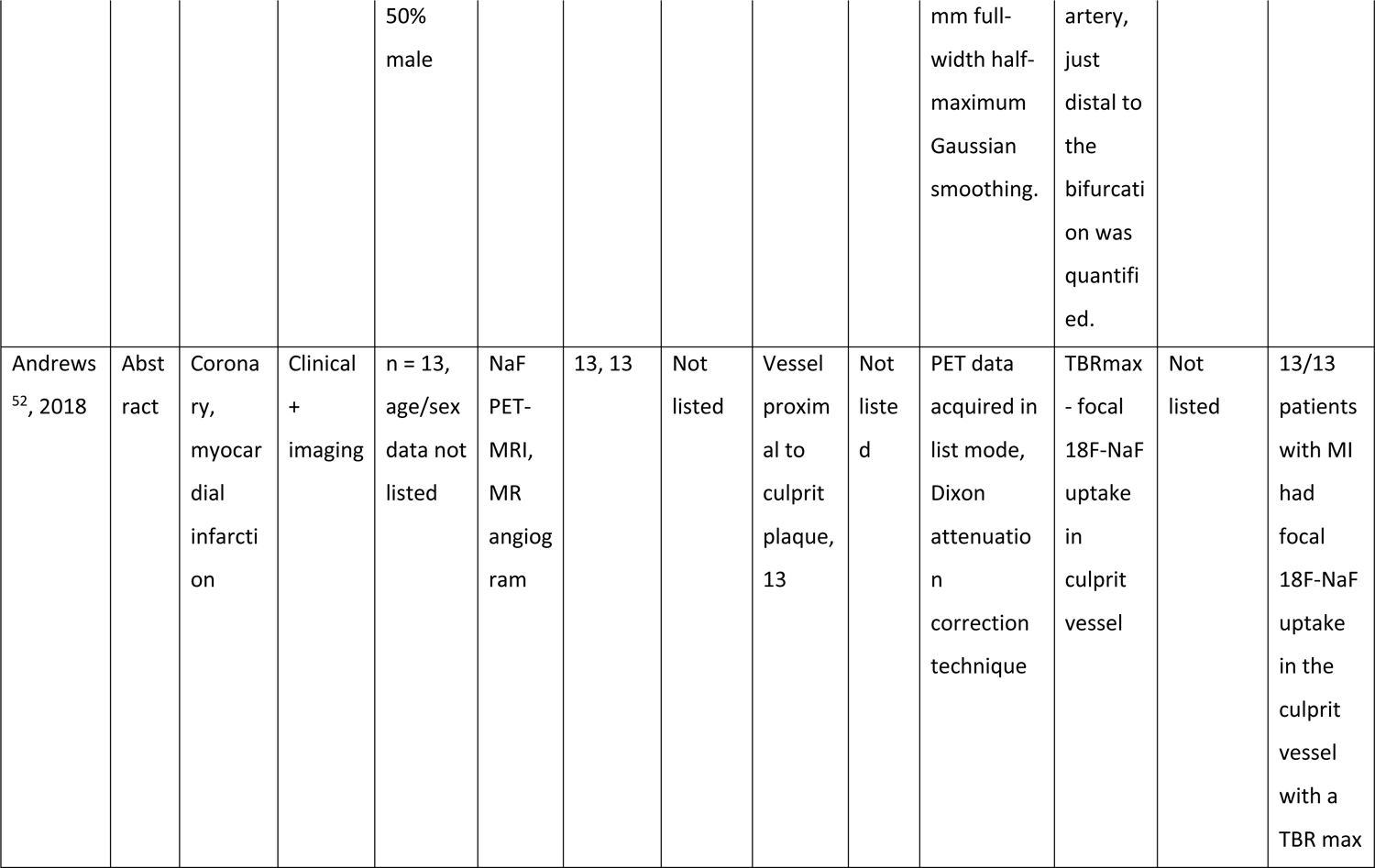
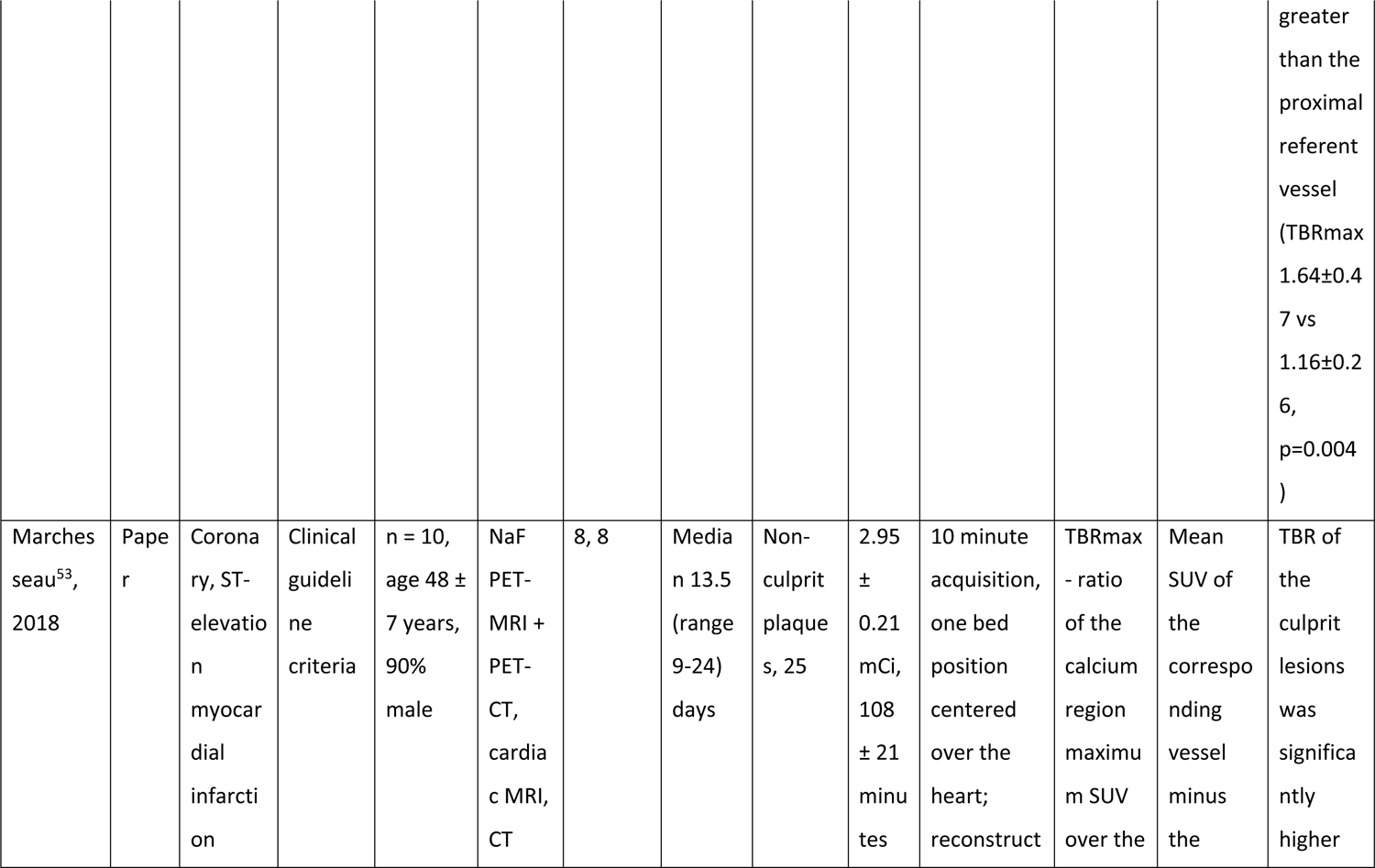
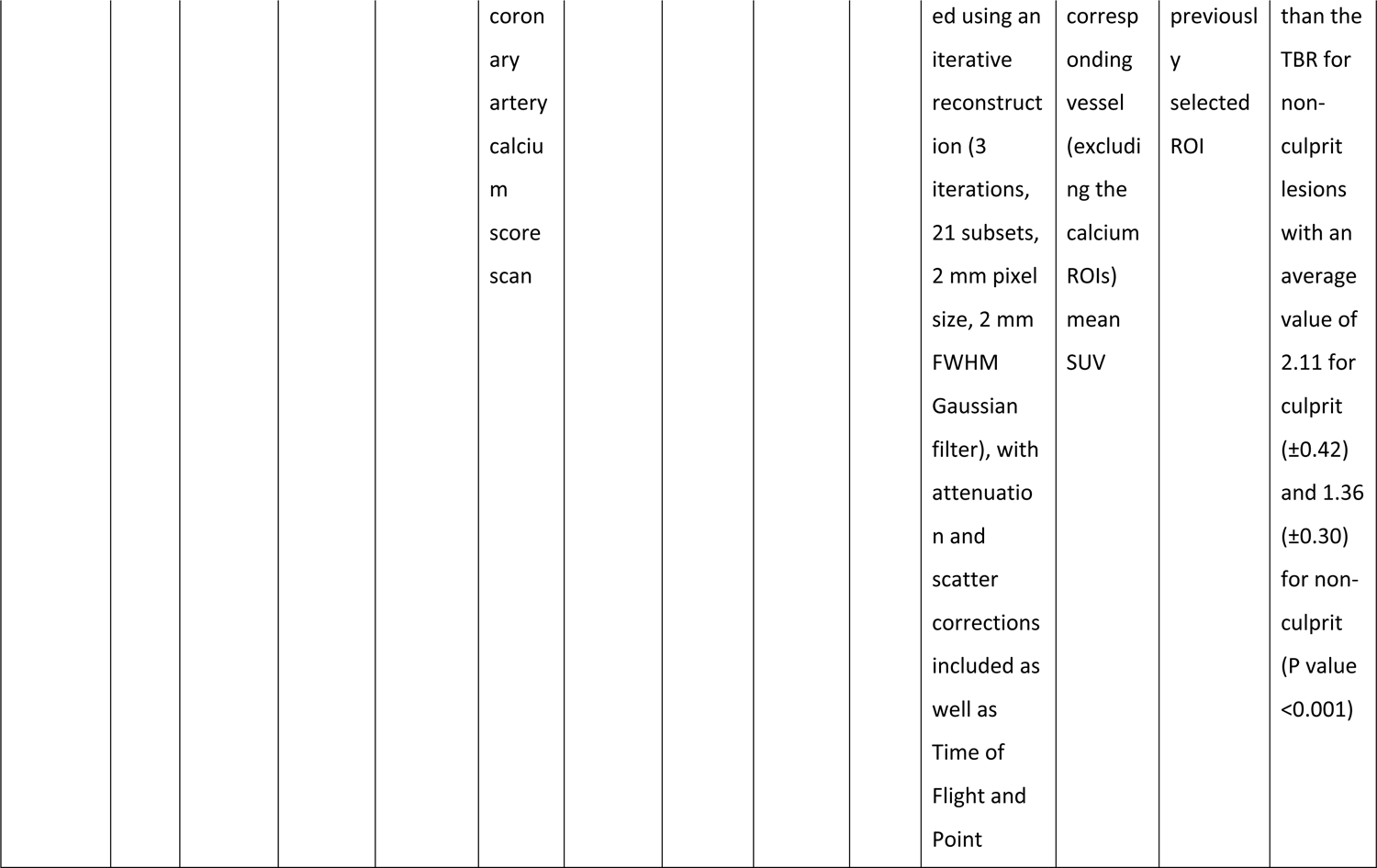
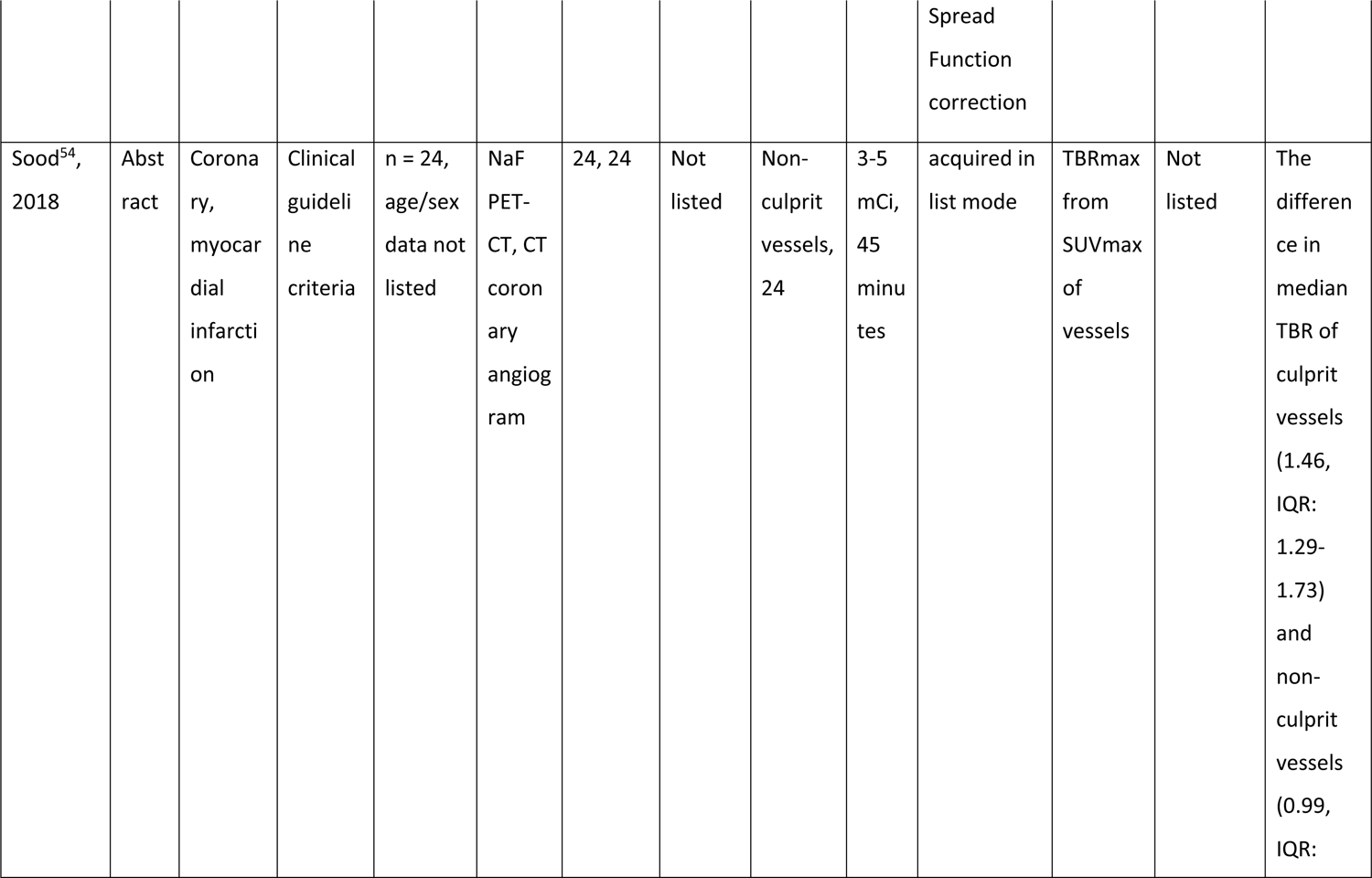
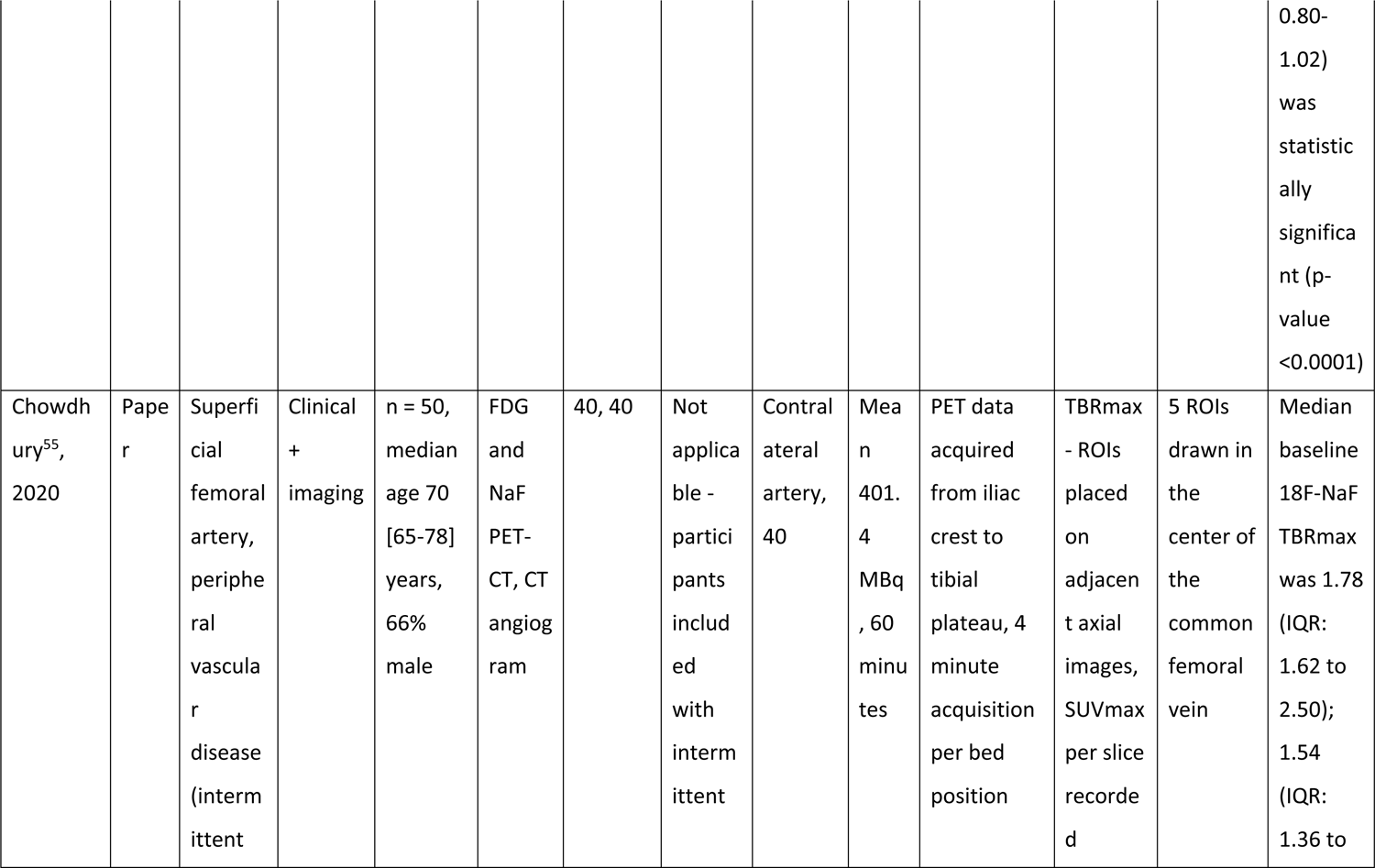
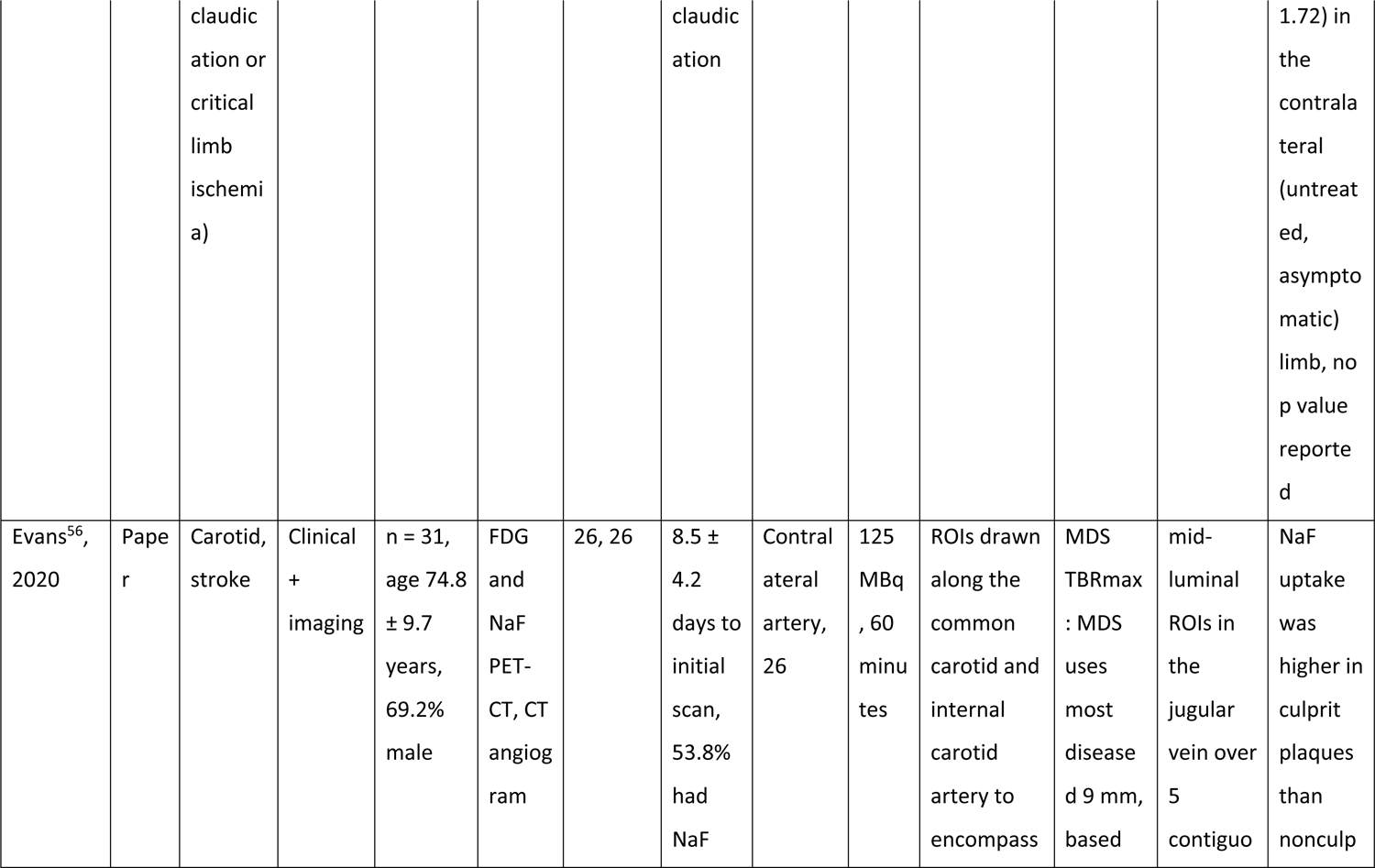
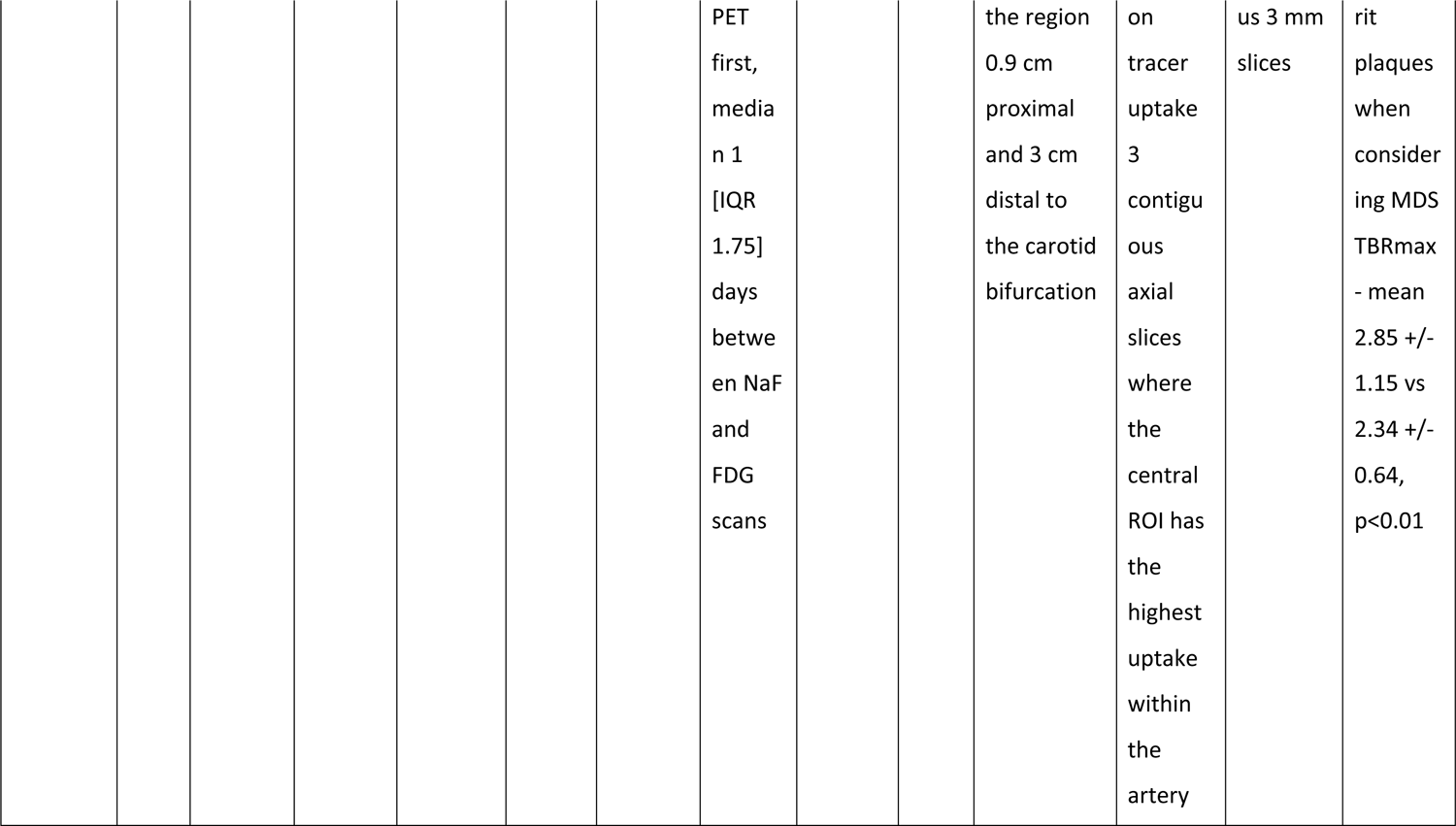
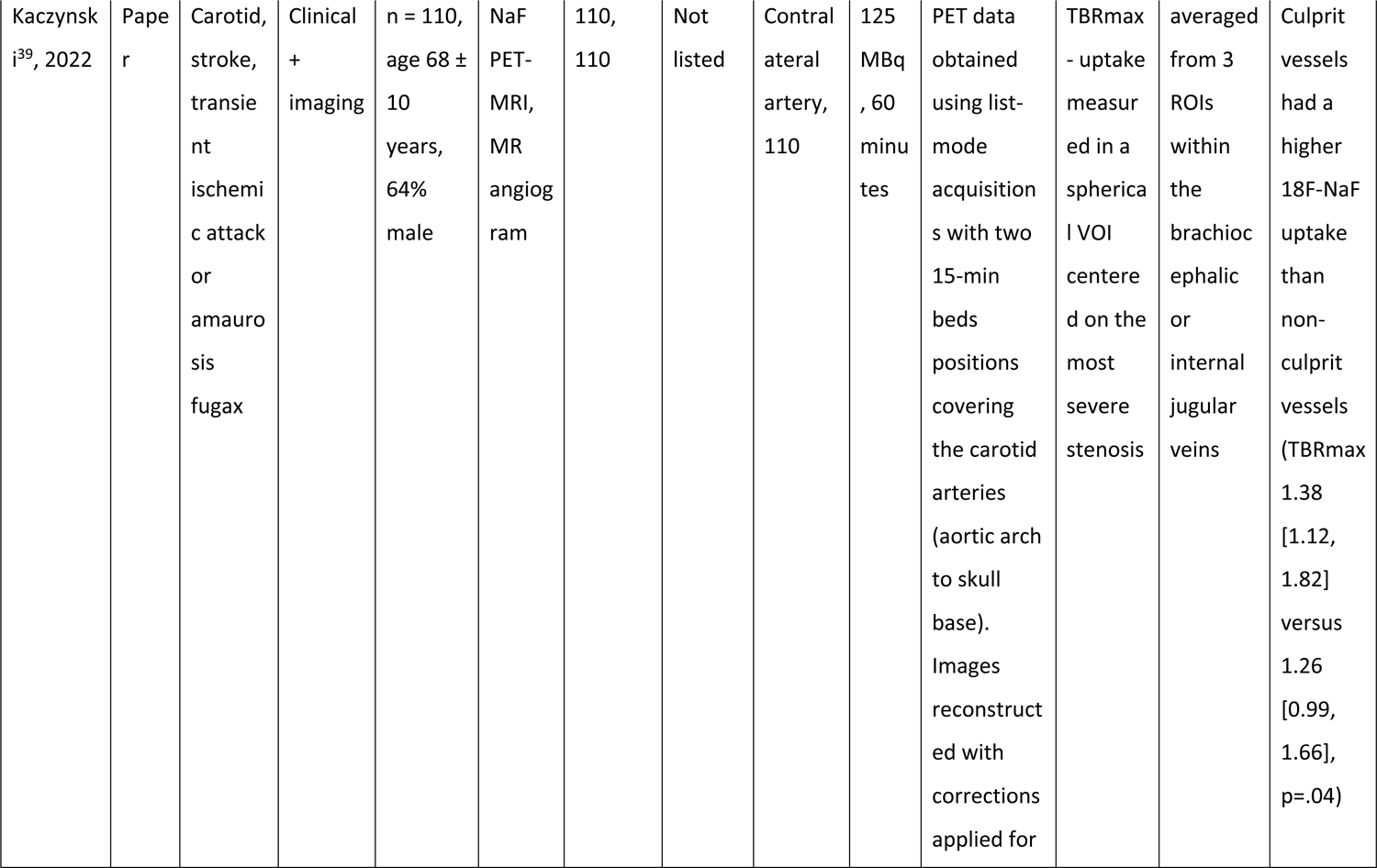
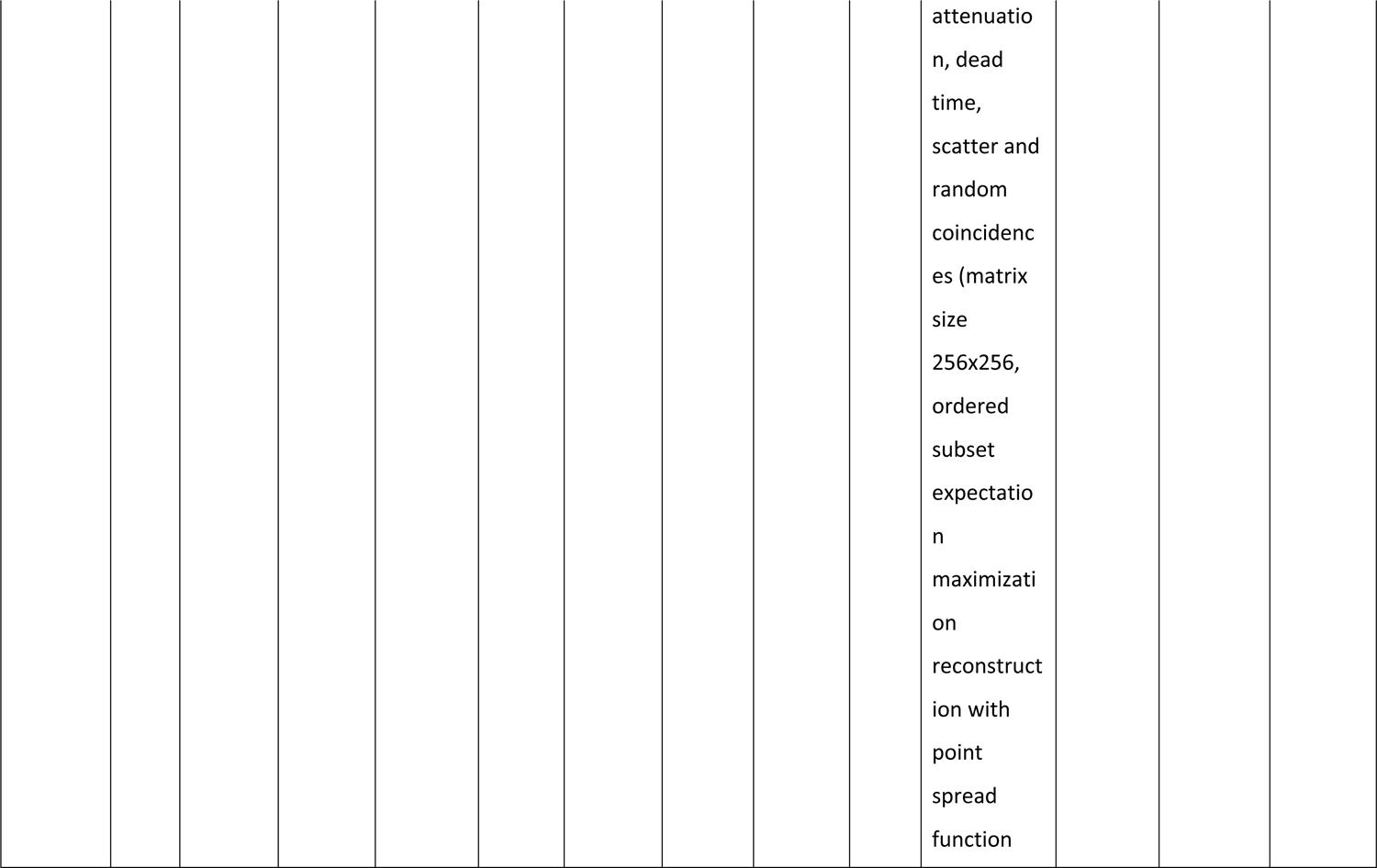
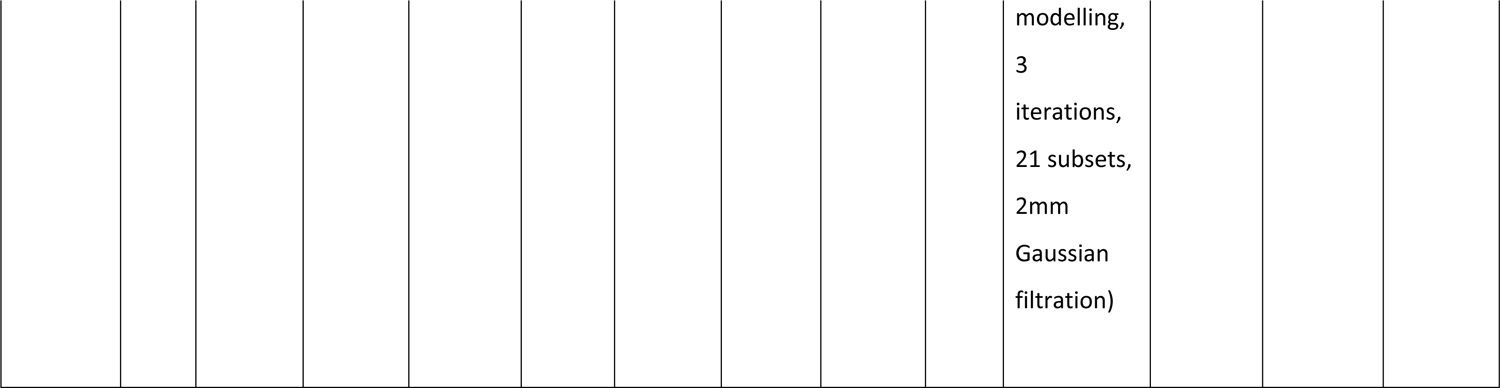
Included study characteristics for studies comparing symptomatic to asymptomatic disease between arteries in the same participant.

**Table 2.**
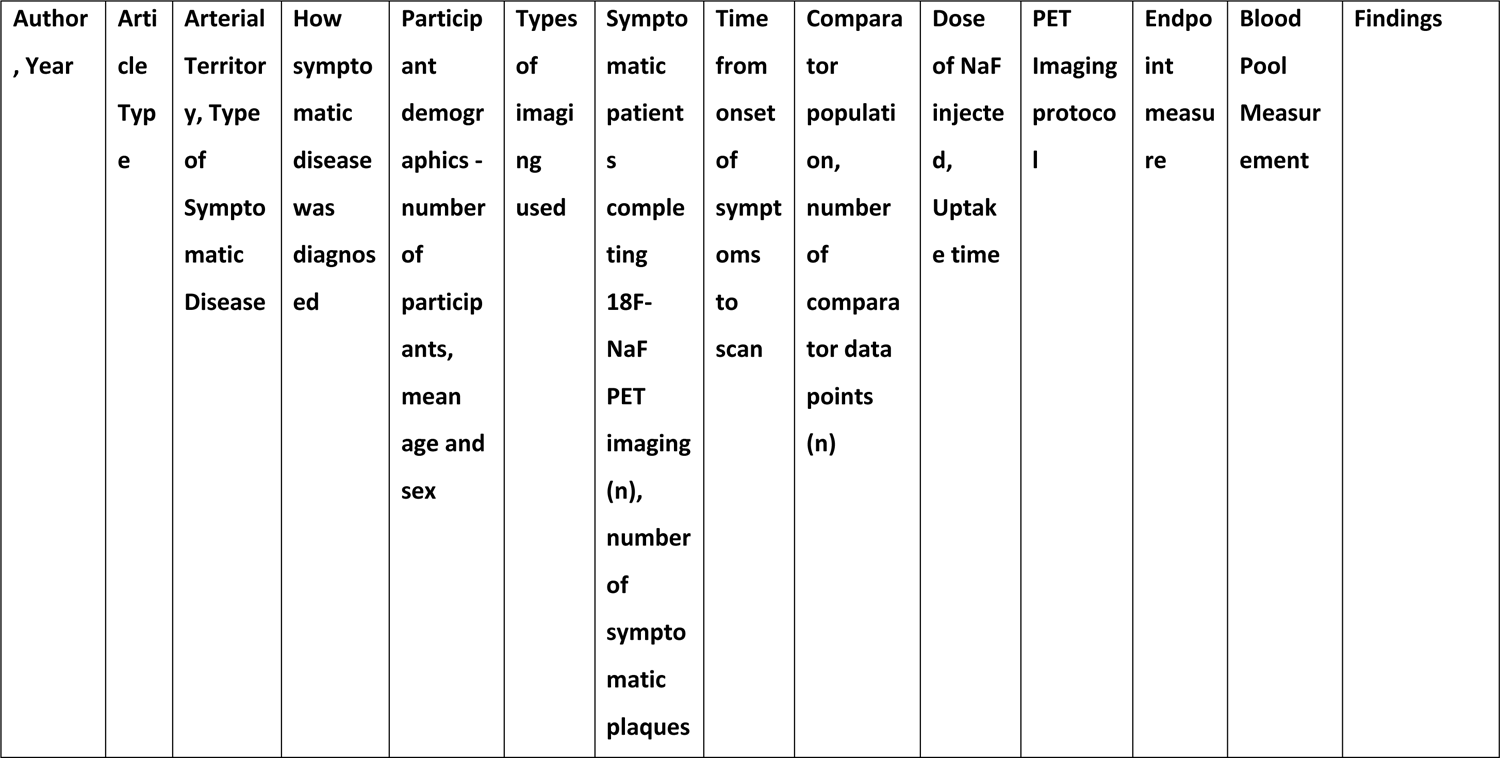
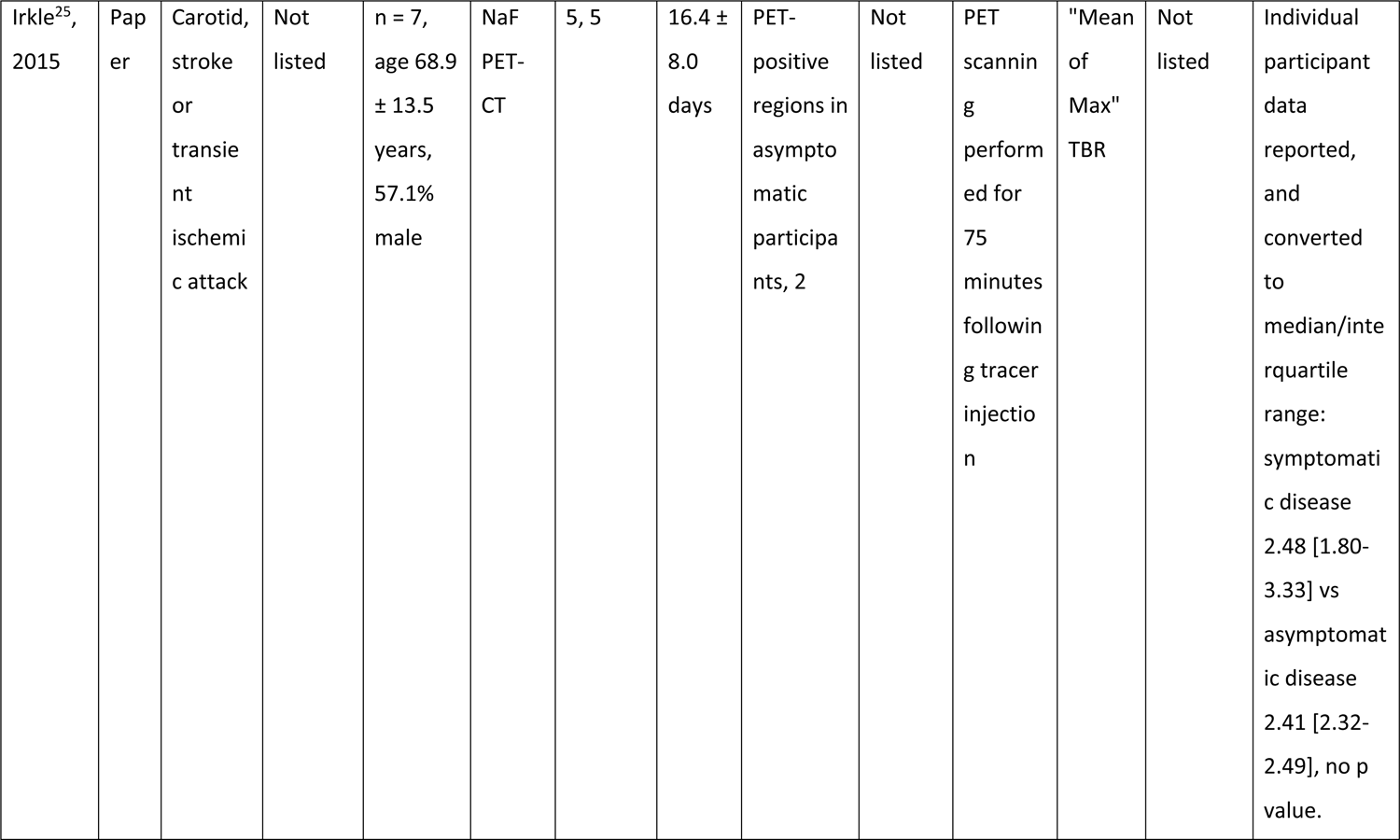
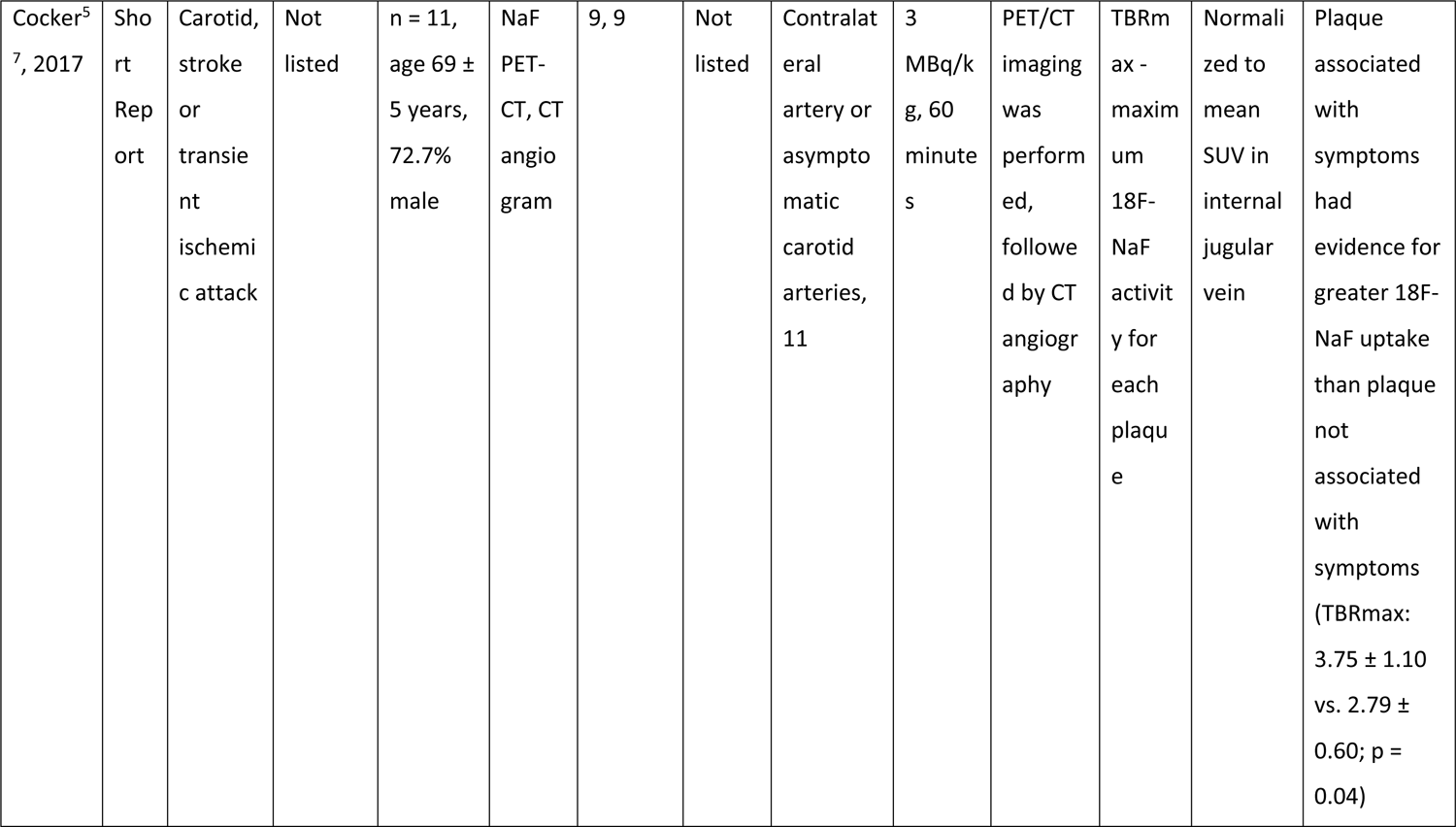
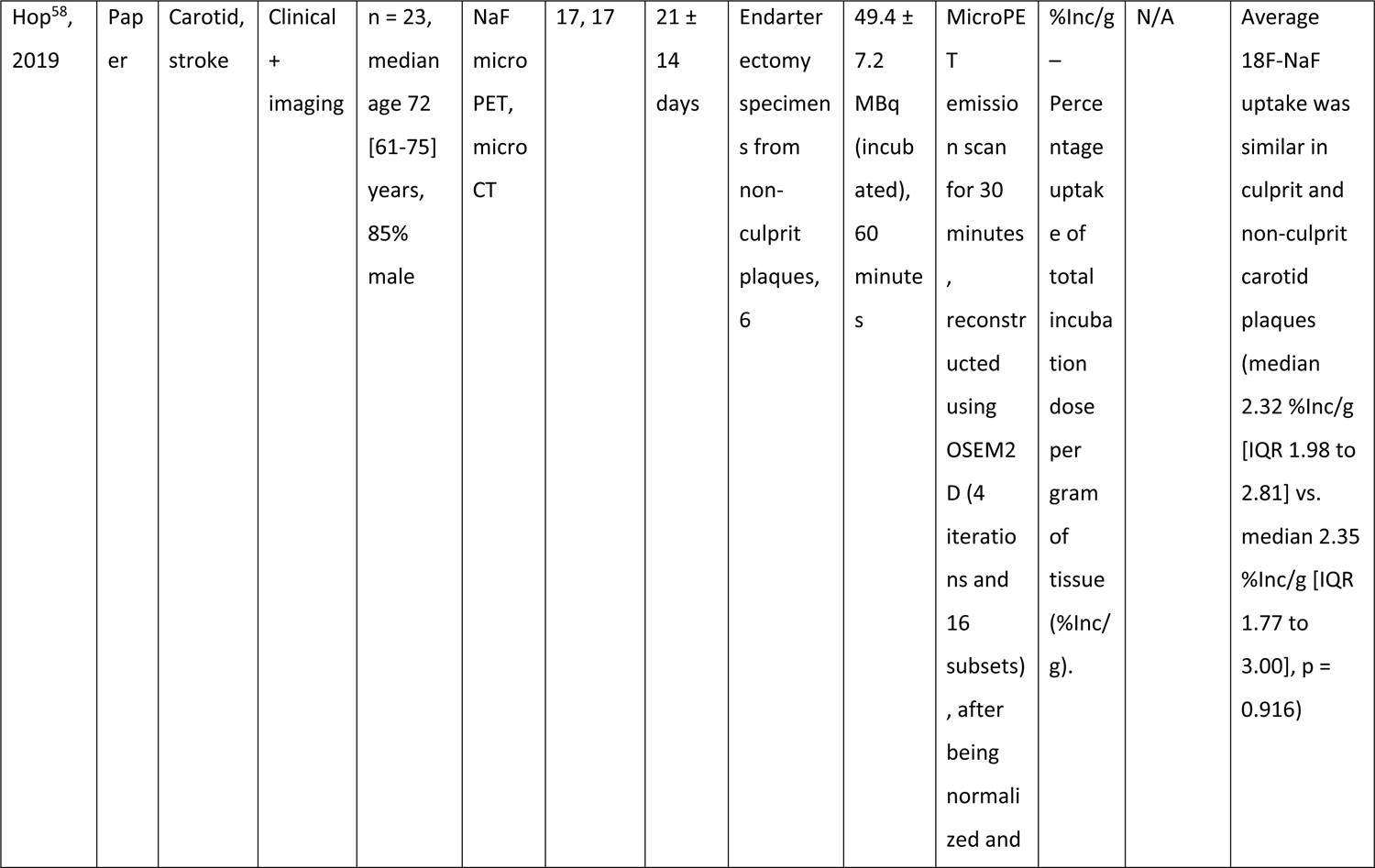
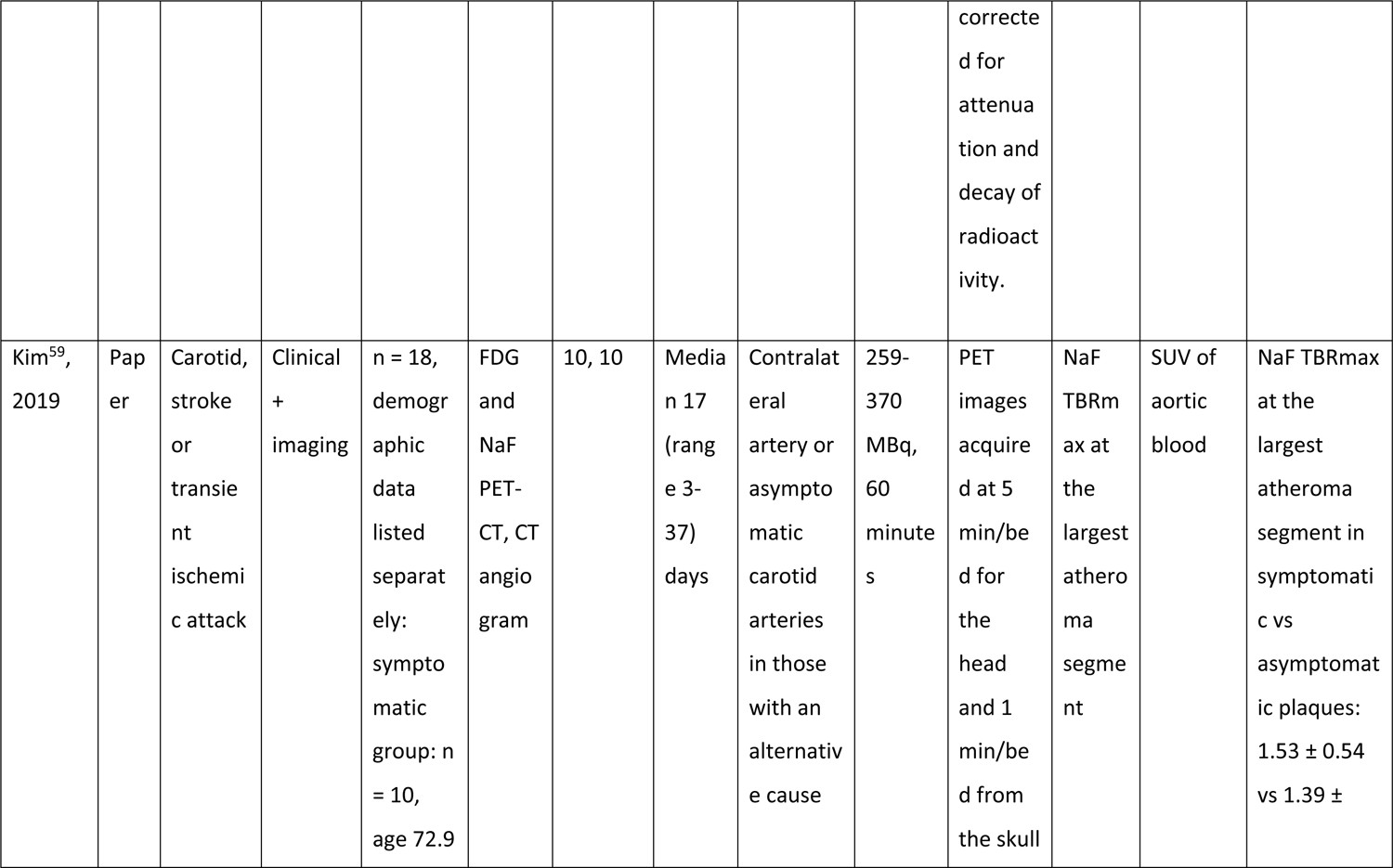
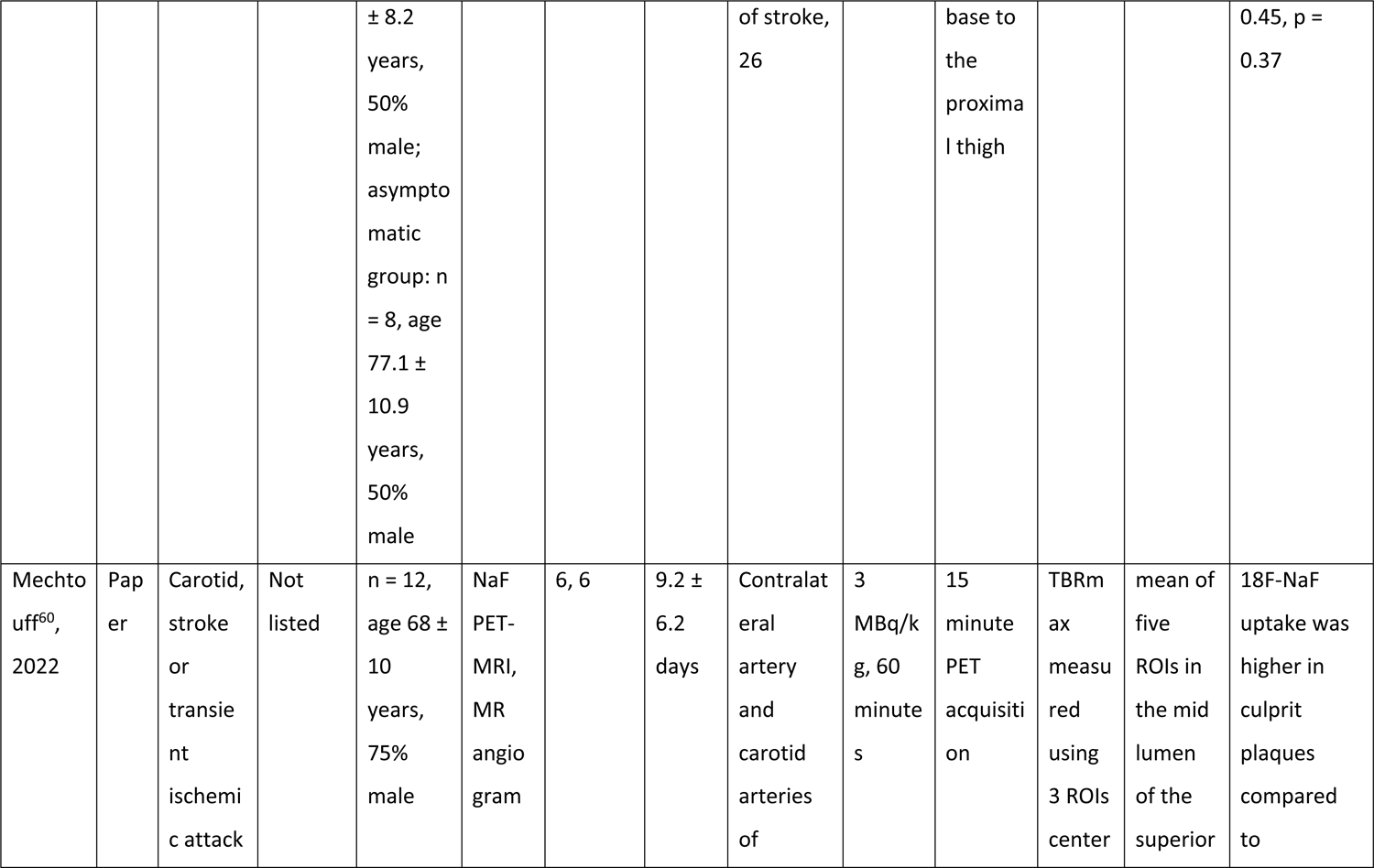
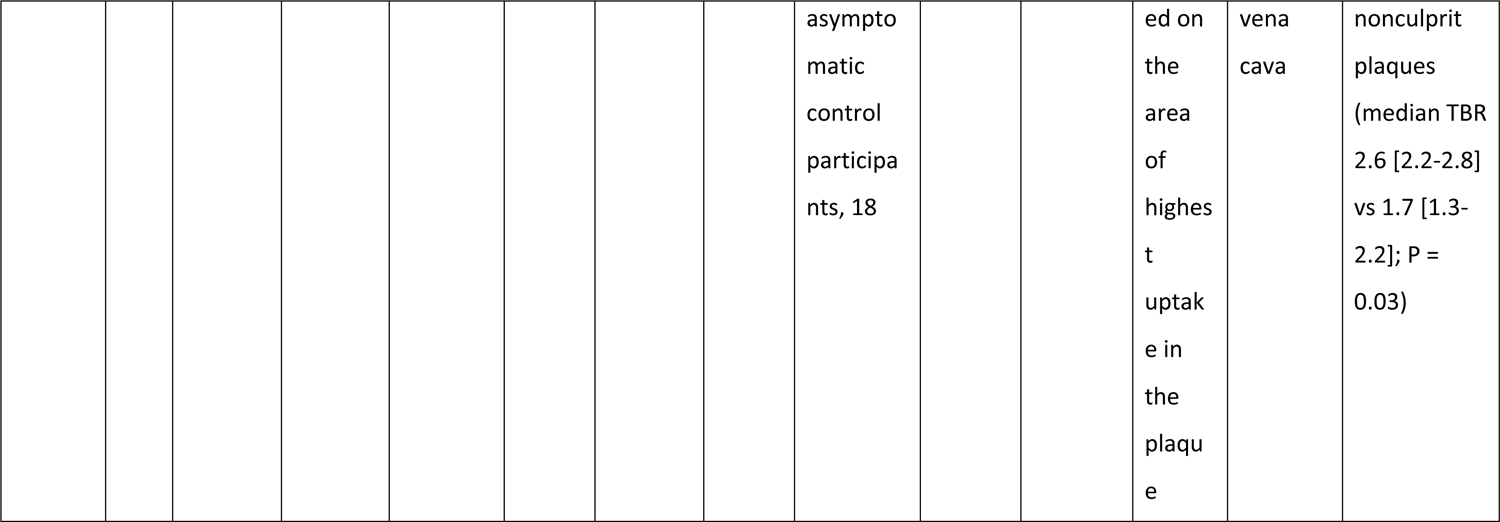
Included study characteristics for studies comparing symptomatic to asymptomatic disease within and between participants.

|  |  |  |  |  |  |  |  |  |  |  |  |  |  |
| --- | --- | --- | --- | --- | --- | --- | --- | --- | --- | --- | --- | --- | --- |
| Irkle<sup>25</sup>,<br>2015 | Pap<br>er | Carotid,<br>stroke<br>or<br>transie<br>nt<br>ischemi<br>c attack | Not<br>listed | n = 7,<br>age 68.9<br>± 13.5<br>years,<br>57.1%<br>male | NaF<br>PET-CT | 5, 5 | 16.4 ±<br>8.0<br>days | PET-positive<br>regions in<br>asympto<br>matic<br>participa<br>nts, 2 | Not<br>listed | PET<br>scannin<br>g<br>perform<br>ed for<br>75<br>minutes<br>followin<br>g tracer<br>injectio<br>n | "Mean<br>of<br>Max"<br>TBR | Not<br>listed | Individual<br>participant<br>data<br>reported,<br>and<br>converted<br>to<br>median/inte<br>rquartile<br>range:<br>symptomati<br>c disease<br>2.48 [1.80-3.33] vs<br>asymptomat<br>ic disease<br>2.41 [2.32-2.49], no p<br>value. |
| Cocker<sup>5</sup><br>7, 2017 | Short<br>Report | Carotid,<br>stroke<br>or<br>transie<br>nt<br>ischemi<br>c attack | Not<br>listed | n = 11,<br>age 69 ±<br>5 years,<br>72.7%<br>male | NaF<br>PET-CT, CT<br>angio<br>gram | 9, 9 | Not<br>listed | Contralat<br>eral<br>artery or<br>asympto<br>matic<br>carotid<br>arteries,<br>11 | 3<br>MBq/k<br>g, 60<br>minute<br>s | PET/CT<br>imaging<br>was<br>perform<br>ed,<br>followe<br>d by CT<br>angiogr<br>aphy | TBRm<br>ax -<br>maxim<br>um<br>18F-NaF<br>activit<br>y for<br>each<br>plaqu<br>e | Normali<br>zed to<br>mean<br>SUV in<br>internal<br>jugular<br>vein | Plaque<br>associated<br>with<br>symptoms<br>had<br>evidence for<br>greater 18F-NaF uptake<br>than plaque<br>not<br>associated<br>with<br>symptoms<br>(TBRmax:<br>3.75 ± 1.10<br>vs. 2.79 ±<br>0.60; p =<br>0.04) |
| Hop<sup>58</sup>,<br>2019 | Paper | Carotid,<br>stroke | Clinical<br>+<br>imaging | n = 23,<br>median<br>age 72<br>[61-75]<br>years,<br>85%<br>male | NaF<br>micro<br>PET,<br>micro<br>CT | 17, 17 | 21 ±<br>14<br>days | Endarter<br>ectomy<br>specimen<br>s from<br>non-culprit<br>plaques,<br>6 | 49.4 ±<br>7.2<br>MBq<br>(incub<br>ated),<br>60<br>minute<br>s | MicroPE<br>T<br>emissio<br>n scan<br>for 30<br>minutes<br>,<br>reconstr<br>ucted<br>using<br>OSEM2<br>D (4<br>iteratio<br>ns and<br>16<br>subsets)<br>, after<br>being<br>normali<br>zed and corrected for attenuation and decay of radioactivity. | %Inc/g<br>–<br>Perce<br>ntage<br>uptak<br>e of<br>total<br>incuba<br>tion<br>dose<br>per<br>gram<br>of<br>tissue<br>(%Inc/<br>g). | N/A | Average<br>18F-NaF<br>uptake was<br>similar in<br>culprit and<br>non-culprit<br>carotid<br>plaques<br>(median<br>2.32 %Inc/g<br>[IQR 1.98 to<br>2.81] vs.<br>median 2.35<br>%Inc/g [IQR<br>1.77 to<br>3.00], p =<br>0.916) |
| Kim<sup>59</sup>, 2019 | Paper | Carotid, stroke or transient ischemic attack | Clinical + imaging | n = 18, demographic data listed separately: symptomatic group: n = 10, age 72.9 | FDG and NaF PET-CT, CT angiogram | 10, 10 | Median 17 (range 3-37) days | Contralateral artery or asymptomatic carotid arteries in those with an alternative cause | 259-370 MBq, 60 minutes | PET images acquired at 5 min/bed for the head and 1 min/bed from the skull | NaF TBRmax at the largest atheroma segment | SUV of aortic blood | NaF TBRmax at the largest atheroma segment in symptomatic vs asymptomatic plaques: 1.53 ± 0.54 vs 1.39 ± |
|  |  |  |  | ± 8.2 years, 50% male; asymptomatic group: n = 8, age 77.1 ± 10.9 years, 50% male |  |  |  | of stroke, 26 |  | base to the proximal thigh |  |  | 0.45, p = 0.37 |
| Mechto<br>uff<sup>60</sup>,<br>2022 | Pap<br>er | Carotid,<br>stroke<br>or<br>transie<br>nt<br>ischemi<br>c attack | Not<br>listed | n = 12, age 68 ± 10 years, 75% male | NaF PET-MRI, MR angiogram | 6, 6 | 9.2 ± 6.2 days | Contralat<br>eral<br>artery<br>and<br>carotid<br>arteries<br>of | 3 MBq/kg, 60 minutes | 15 minute PET acquisition | TBRmax measured using 3 ROIs center | mean of five ROIs in the mid lumen of the superior | 18F-NaF uptake was higher in culprit plaques compared to |
|  |  |  |  |  |  |  |  | asymptomatic<br>control<br>participants, 18 |  |  | ed on<br>the<br>area<br>of<br>highest<br>uptake<br>in<br>the<br>plaque | vena<br>cava | nonculprit<br>plaques<br>(median TBR<br>2.6 [2.2-2.8]<br>vs 1.7 [1.3-2.2]; P =<br>0.03) |

Subgroup analysis was performed between the different arterial territories assessed in the included studies to generate standardized mean difference measurements for symptomatic atherosclerotic plaques compared with asymptomatic plaques, and to determine any statistical differences in NaF uptake between different arterial territories.

## Results

### Included Studies

A total of 966 titles were initially identified from the search (**Figure 4**)^37^. Manual de-duplication of results was performed, and the remaining 733 records underwent manual screening of the titles and abstracts^38^. Of those, 636 were excluded as not meeting the inclusion criteria. 97 articles were therefore sought for retrieval. Five articles were not available for analysis and 92 articles were therefore assessed for eligibility through full-text review. Of these, 77 were excluded, 53 due to not reporting outcomes related to symptomatic atherosclerotic disease, 15 due to no documented comparison between symptomatic and asymptomatic disease, eight due to discussion of imaging protocols and quality assessment and one due to reporting outcome measures insufficient for meta-analysis. The remaining 15 articles were included in the meta-analysis, and form the study population analyzed. A summary of study details is shown in **Tables 1 and 2**, dichotomized based on the comparator of symptomatic disease used in each study.

**Figure 4.**
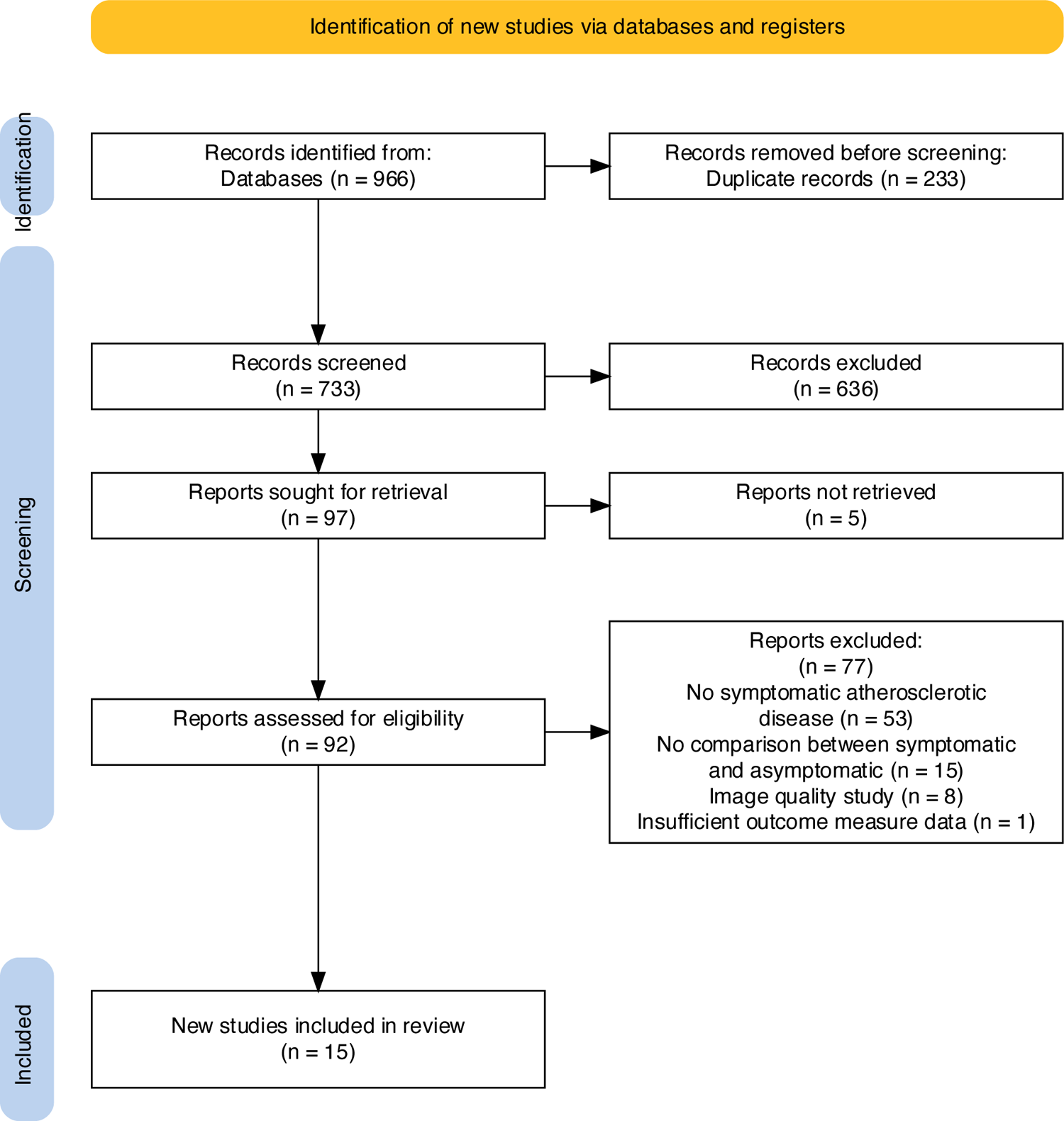
PRISMA diagram of systema7c review search synthesis

Comparative intra-individual ^18^F-NaF tracer uptake between symptomatic and asymptomatic atherosclerotic disease Ten studies, including 352 participants, reported data in participants comparing NaF uptake in the symptomatic atherosclerotic plaque compared to asymptomatic plaques within the same participant’s vascular territory being observed, including data for 317 symptomatic, and 336 asymptomatic plaques (**Table 1**).

Pooled comparisons of these studies demonstrated a significantly higher uptake in symptomatic lesions compared to asymptomatic plaques (standardized mean difference 0.42, 95% CI 0.29-0.56, *p*<0.001, **Figure 5**). Subgroup analysis to compare results from different arterial territories showed no significant difference in this relationship based on the site of the symptomatic plaque (Q_M_ = 5.02, *p* = 0.08). Significant heterogeneity was noted between the studies (Q = 19.89, I^2^ = 54.1%, p = 0.019).

**Figure 5.**
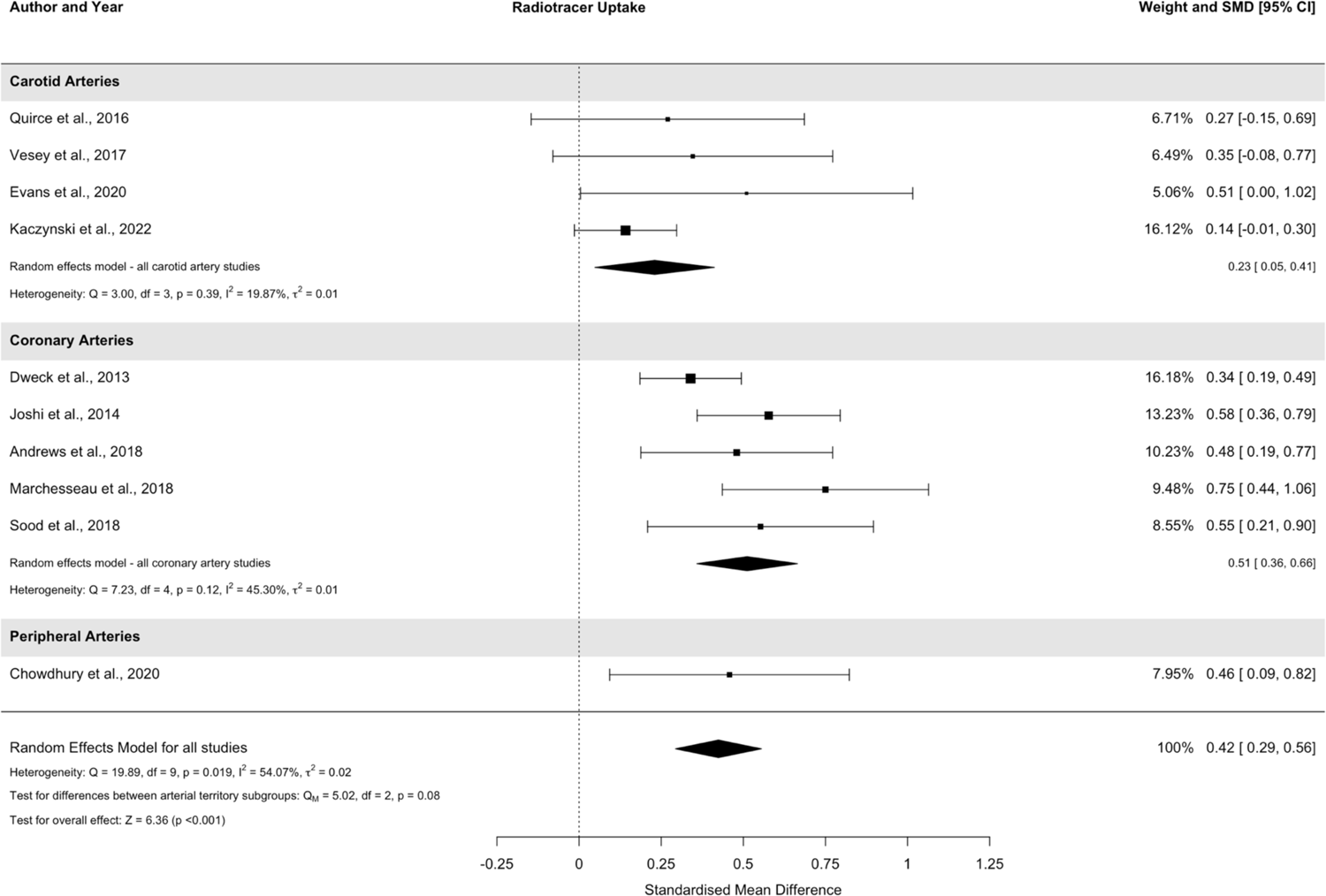
Forest Plot of included studies summarising data comparing symptomatic and asymptomatic atherosclerotic disease within individuals. SMD = standardized mean difference, CI = confidence intervals

### Comparative NaF tracer uptake in symptomatic and asymptomatic atherosclerotic plaques where a healthy control population was included

Five studies, including 71 participants, reported data in participants comparing NaF uptake in the symptomatic atherosclerotic plaque compared to asymptomatic plaques, pooling data from asymptomatic disease within the same symptomatic individual and plaques from asymptomatic healthy controls, including data for 47 symptomatic, and 63 asymptomatic plaques (**Table 2**).

Analysis of the pooled data from these studies demonstrated a significantly higher NaF uptake in symptomatic atherosclerotic lesions compared to asymptomatic lesions, present in the same individuals, or in healthy controls (standardized mean difference 0.44, 95% CI 0.03-0.85, *p* = 0.037, **Figure 6**). All studies analyzed assessed atherosclerotic disease within the carotid arteries, and subgroup analysis for differences between different arterial territories was therefore not conducted. There was no significant heterogeneity between studies included in this analysis (Q = 6.37, I^2^ = 40.4%, p = 0.17).

**Figure 6.**
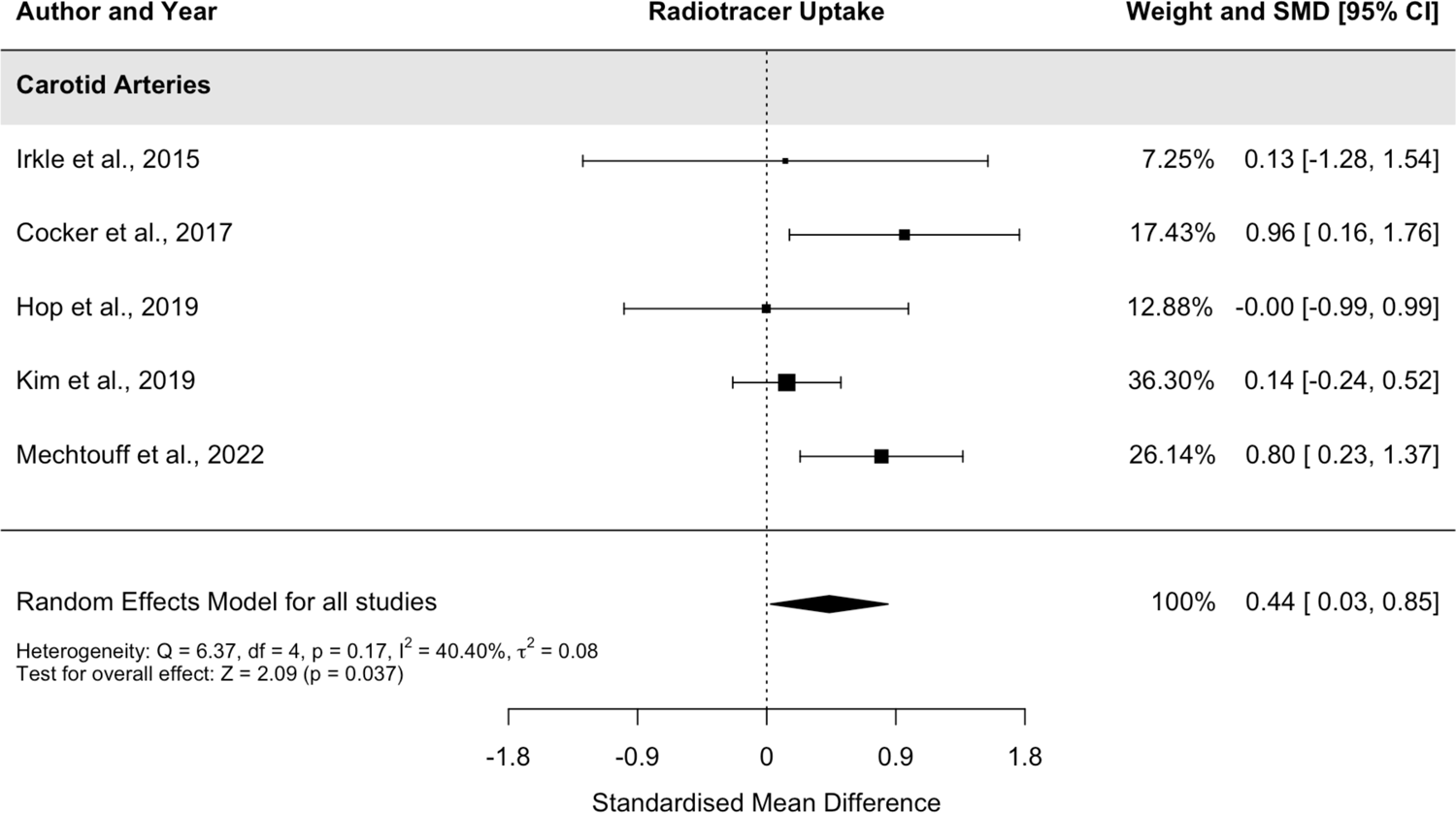
Forest Plot of included studies summarizing data comparing symptomatic and asymptomatic atherosclerotic disease within individuals. SMD = standardized mean difference, CI = confidence intervals

## Discussion

These results demonstrate the utility of NaF-PET, combined with CT or MRI, to differentiate between symptomatic and asymptomatic atherosclerotic disease in a number of different vascular territories. Studies have also demonstrated a correlation between NaF uptake and high-risk morphological features on MRI, such as the presence of a lipid-rich necrotic core, or intraplaque haemorrhage^39^. In addition, there is emerging evidence of a prognostic link between NaF signal on PET and the risk of recurrent disease^40^, where coronary NaF imaging had the ability to predict myocardial infarction and cardiovascular death.

As noted in **Tables 1 and 2**, there are a range of different doses of NaF used, along with a non-standardized uptake time, blood pool measurement for TBR calculations, and outcome measurement. Irkle et al^25^ demonstrated an optimum uptake time of around 60 minutes, based on *in vitro* and *in vivo* data. However, NaF-PET lacks a consensus on best practice, in contrast to vascular FDG-PET imaging, following the 2005 position paper from the European Association of Nuclear Medicine^41^. Having a standardized methodology for performing and analyzing vascular NaF imaging would allow increased comparability and reproducibility between studies.

There was also significant heterogeneity in the included studies between the time of symptom onset and imaging being performed. There is little data concerning the temporal changes in microcalcification and NaF uptake following an acute atherosclerotic event, and further information on the optimum time period for imaging microcalcification in relation to symptomatic disease could further standardize imaging protocols and improve robustness of outcomes measured using NaF-PET as a surrogate marker.

In atherosclerosis, PET has been demonstrated to have a high sensitivity for the target pathophysiology and can be combined with other imaging modalities to provide additional utility, such as in combination with CT and CT angiography to assess for stenotic disease^42^ and calcium scores in the coronary vasculature^43^, or with MRI, to identify high risk appearances as discussed above^44^. In vascular imaging, NaF may be superior to FDG as it may have a superior ability to discriminate between symptomatic and asymptomatic disease in high-risk individuals^45^. In addition, NaF does not require fasting prior to the uptake period, and can be used with a shorter uptake time compared to FDG-PET^25,41^. NaF is also less susceptible to spill-over artefact, such as from the myocardium, which can limit FDG-based PET imaging of the arteries^45^. However, the cost, radiation exposure and uptake and scanning time mean the role of PET imaging in routine clinical assessment of atherosclerotic disease is currently limited.

Microcalcification is known to confer an increased risk of plaque rupture through enzymatic^18^ and mechanical^17^ destabilization of the plaque surface. Therefore, targeting this process may reduce the risk of early recurrence following symptomatic atherosclerotic disease. NaF-PET can be utilized to assess responses to clinical interventions, given its accuracy and sensitivity in determining differences in the presence of microcalcification *in vivo*^46^. Additionally, the process by which microcalcification evolves into macrocalcification, which is thought to be protective for the atherosclerotic plaque, is poorly defined and understood^47^. Temporal evaluation of the microcalcification-macrocalcification process through NaF-PET/CT could shed more light on the factors which confer a greater or lesser degree of risk with calcification in atherosclerosis. In addition, NaF imaging could potentially have a role in risk stratification in patients where there is uncertainty about the risk of recurrent stroke or the need for surgical intervention, which is particularly relevant for those with moderate 50-69% stenoses.

## Conclusion

Our findings support vascular NaF-PET imaging as a reliable method for assessment of atherosclerotic disease that is able to help differentiate between symptomatic/vulnerable plaques and asymptomatic/stable plaques. The majority of data involves analysis of the carotid or coronary circulation, but NaF-PET imaging is also a viable imaging technique in other arterial territories. There is a potential future role of NaF-PET in clinical atherosclerosis imaging and providing surrogate markers of impact in interventional trials, but harmonization of imaging protocols including outcome measures, injected doses and uptake times is required to ensure comparability between studies.

## Funding

SB is supported by a Research Training Fellowship from The Dunhill Medical Trust [JBGS22\20]. The ICARUSS study was supported as part of a Research Training Fellowship awarded to NRE by The Dunhill Medical Trust [RTF44/0114], and by the NIHR Cambridge Biomedical Research Centre (NIHR203312). MC is supported by a BHF Career Development Fellowship and the NIHR. JMT is supported by a Wellcome Trust Clinical Research Career Development Fellowship (211100/Z/18/Z) and the Cambridge BHF Centre for Research Excellence (18/1/34212). JHFR is part-supported by the NIHR Cambridge Biomedical Research Centre, the British Heart Foundation, HEFCE, the EPSRC and the Wellcome Trust. NRE is supported by a Stroke Association Senior Clinical Lectureship [SA-SCL-MED-22\100006].

## Disclosures

## Non-standard Abbreviations and Acronyms

MDS: most diseased segment

NaF: sodium fluoride

ROI: region of interest

SUV: standardized uptake value

TBR: tissue-to-background ratio

TBRmax: maximum tissue-to-background ratio from the region of interest

## Data Availability

All anonymised data will be provided on reasonable request to the corresponding author.

---

**Supplementary Figure 1.**
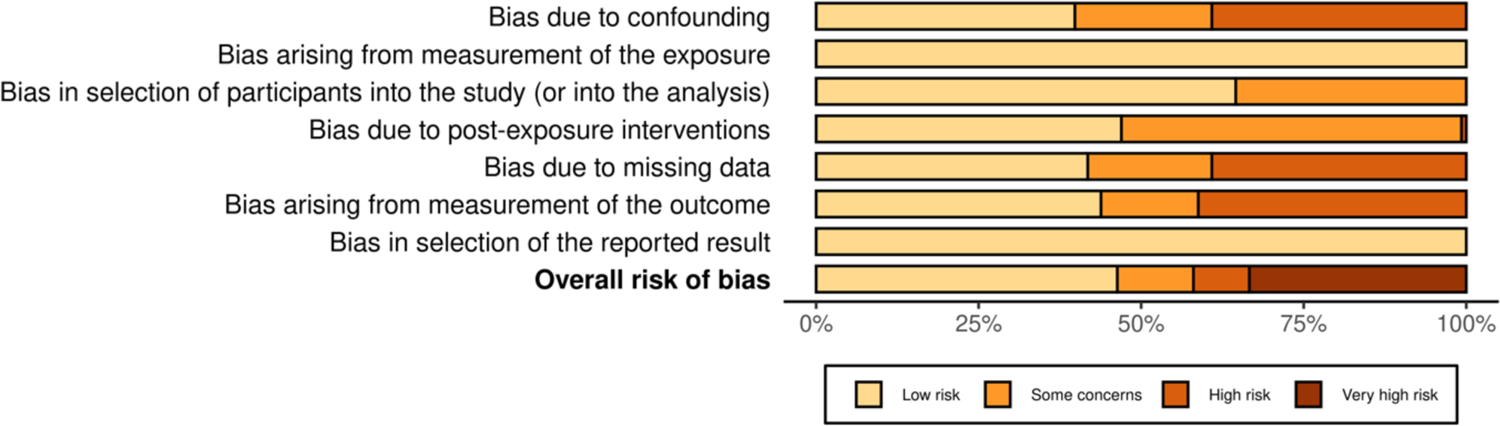
Summary plot of Risk of Bias assessment using ROBINS-E (Risk Of Bias In Non-randomized Studies - of Exposures) tool

## Notes

### Competing Interest Statement

The authors have declared no competing interest.

### Author Declarations

Not applicable Study registered to PROSPERO: www.crd.york.ac.uk/CRDWeb/ShowRecord.asp?ID=42023451363

